# Accurate repeat expansion genotyping from whole-genome and real-time targeted nanopore sequencing with Mahi-mahi

**DOI:** 10.64898/2026.10.16.26365781

**Authors:** Sophia B. Gibson, Rebecca N. Palmer, Sebastian Hollizeck, Wouter Koole, Heather M. Jeffery, Mirjan Albring, Jillian G. Buchan, R. Keira Cheetham, Martin G. Elferink, Joy Goffena, Elizabeth J. Hamlin, Rumo Jansen, Paul J. Lockhart, Rachel Martin, Angela L. Miller, Inge van Outersterp, Trent Prall, Jenna Rosinski, Simone M. Rowley, Simon Sadedin, Danielle Sewell, Macgregor Todd, Alexey Zabelkin, Miranda P.G. Zalusky, Bert van der Zwaag, Sergey Nurk, Sissel Juul, Koen L.I. van Gassen, Sebastian Lunke, Danny E. Miller, Philipp Rescheneder

## Abstract

Tandem repeat expansions cause more than 60 Mendelian disorders but are difficult to detect and size accurately with short-read sequencing because of their length and repetitive structure. Long-read sequencing (LRS) resolves these loci directly, yet analytic tooling for nanopore sequencing data has lagged behind that available for other long-read platforms. Here we present Mahi-mahi, a tandem repeat genotyper designed for nanopore reads that reports allele-level repeat length, motif composition, interruptions, and read-level flags for systematic basecalling artifacts. Across six Genome in a Bottle samples, whole-genome and adaptive sampling (real-time targeted) data returned calls within one repeat unit of each other for 829 of 832 alleles, and Mahi-mahi calls were within one repeat unit of orthogonal HiFi data at 95.6% of compared alleles after excluding three loci at which the two tools define the repeat differently. In 245 samples from the 1000 Genomes Project, Mahi-mahi calls were within one repeat unit of assembly-derived calls for 98.6% of 10,927 alleles at 57 loci, and Mahi-mahi distinguished pathogenic from nonpathogenic *RFC1* alleles of overlapping size by motif. Across 14 Coriell reference lines, Mahi-mahi identified the expected expansion in every line and agreed with orthogonal assays to within two repeat units at *HTT*, *ATXN1*, *DMPK*, and *PABPN1*. Finally, in 103 samples from three laboratories, Mahi-mahi detected an expanded allele in all samples in which prior testing had reported a pathogenic expansion, assigned the same category as the referring laboratory for 98% of 180 genotypes, and identified known *ATXN1* interruption structure. Our results demonstrate that Mahi-mahi accurately identifies repeat expansions at disease-associated loci and could serve as a single assay to evaluate repeat expansions.

## INTRODUCTION

Tandem repeat expansions are a mechanistically distinct and clinically important class of variation, causing more than 60 Mendelian conditions spanning neurological, neuromuscular, and developmental phenotypes (Hiatt et al. 2025; Depienne and Mandel 2021). They are systematically missed by exome sequencing and, to a large extent, by short-read genome sequencing as pathogenic alleles may exceed read and fragment length, GC-rich and structurally complex motifs may sequence poorly, and pathogenic alleles may be defined by motif composition or interruption structure rather than by length alone. As a result, repeat expansion testing is often performed locus-by-locus with tests such as repeat-primed PCR, Southern blot, or optical genome mapping ordered one gene at a time, which leads to increased testing costs, prolonged turnaround times, and extended diagnostic workups.

Long-read sequencing (LRS) is unique as longer reads are able to completely span most pathogenic alleles allowing simultaneous identification of repeat length and sequence rather than inferring size from flanking evidence (Miller et al. 2021; Stevanovski et al. 2022). A large number of tandem repeat callers have been published for nanopore data, including tandem-genotypes, Straglr, NanoRepeat, and STRdust, alongside platform-agnostic tools such as LongTR, vamos, STRkit, and ATaRVa (Mitsuhashi et al. 2019; Fang et al. 2023; De Coster et al. 2024; Ziaei Jam et al. 2024; Ren et al. 2023; Lougheed et al. 2026; Sivakumar et al. 2025; Readman et al. 2021). However, these tools differ substantially in their locus catalogs, output formats, and the repeat features they report; and few were designed around the error characteristics of ONT reads at repetitive sequence. In contrast, analysis of PacBio HiFi data has largely converged on a single tool, TRGT (Dolzhenko et al. 2024), which provides sequence-resolved, catalog-driven genotyping across clinically relevant loci. A similar consensus exists for short-read data, where ExpansionHunter and ExpansionHunter Denovo are commonly used (Dolzhenko et al. 2017, 2020). Thus, despite offering capabilities that neither short-read nor HiFi sequencing provide, such as no upper limit on read length, and adaptive sampling for target enrichment (Loose et al. 2016), ONT sequencing lacks a single disease-oriented tandem repeat genotyper.

To address this gap, we developed Mahi-mahi (https://github.com/nanoporetech/mahi-mahi), a tandem repeat genotyper designed specifically for nanopore data. Mahi-mahi uses a curated locus catalog to extract reads spanning each target, decomposes the repeat structure of every read into its constituent motifs, clusters reads into alleles, and reports allele-level repeat length, motif composition, and interruption structure together with read-level quality flags. Read-level flags address the problem that basecalling error at tandem repeats is not random, and certain motifs generate systematic, strand-biased miscalls across several reads within a locus (Wick et al. 2019). Because these errors are consistent rather than stochastic, they are not removed by consensus approaches and can produce a confidently incorrect motif call even when repeat length is unaffected. Mahi-mahi identifies reads affected by basecalling errors, flags and excludes them from motif analysis, but preserves them for repeat length analysis, thus enabling accurate genotyping and repeat length sizing across TR loci.

Here, we evaluate Mahi-mahi across multiple cohorts chosen to interrogate different aspects of performance using both whole-genome and adaptive sampling with a targeted catalog of genes known to harbor disease-causing tandem repeat expansions. In six Genome in a Bottle (GIAB) samples (Zook et al. 2016) sequenced by both whole-genome and adaptive sampling, the two strategies returned calls within one repeat unit of each other for more than 99% of alleles, with high concordance with repeat lengths identified by orthogonal LRS approaches. In 245 available samples sequenced on the R10 flow cell from the 1000 Genomes Project, Mahi-mahi calls were within one repeat unit of assembly-derived genotypes for 98.6% of alleles, and Mahi-mahi recovered every previously reported expansion supported by the sequencing reads as well as five expanded alleles not identified using an assembly-based approach (Gustafson et al. 2024; Gibson et al. 2026). Among 14 Coriell reference lines, Mahi-mahi identified the expected expansion in every line, and across 103 samples from three laboratories with orthogonal genotypes, Mahi-mahi detected an expanded allele in every sample with a pathogenic expansion reported by prior testing and matched the reported allele size within one repeat unit for 90% of alleles below 100 repeat units. Thus, Mahi-mahi provides sequence-resolved tandem repeat genotyping from nanopore data at a high level of precision, and is compatible with both whole-genome and targeted sequencing strategies.

## METHODS

### Mahi-mahi algorithm and implementation

Mahi-mahi characterizes tandem repeat length, motif composition, interruptions, and CpG modification directly from reference-aligned nanopore sequencing reads **(Supplemental Figure 1)**. It first retrieves reads overlapping each locus in the input BAM and uses reference-flank realignment to refine repeat boundaries and to recover spanning reads from misaligned or soft-clipped alignments. Extracted repeat sequences are then decomposed using TRviz (Park et al. 2023), retaining motif order and counts for each read. Strand-specific differences in motif-derived k-mer frequencies identify potentially erroneous reads. By default, flagged reads are excluded from motif and CpG modification summaries while their length evidence is retained.

Allele inference combines length clustering with selective use of informative non-spanning reads and existing haplotags. With at least two spanning reads per cluster, preliminary clustering estimates the longest supported allele, falling back to the median spanning-read length when clustering is inconclusive. Non-spanning reads enter length estimation only when they reach this estimate plus a margin of 75 bp by default; with fewer spanning reads, this cutoff is zero. Repeat lengths from spanning and informative non-spanning reads are clustered by kernel density estimation with a length-dependent bandwidth, peak-support and separation criteria, and rules accommodating broad distributions. An alternative bandwidth is tested when clustering is unsuccessful or ambiguous; motif decompositions provide an additional check for closely spaced candidate alleles. Read haplotype information is used to resolve overlapping length distributions or provide a fallback when clustering is unresolved, whereas confident clustering takes precedence over conflicting read haplotags. Composite loci are clustered jointly before their decomposed sequences are partitioned into components, preserving allele assignments. At composite loci, sections are ordered and each must form a contiguous block, so that a section’s count includes any interruptions within it. Allele lengths are reported as the median of the repeat lengths contributing to a given cluster, or the upper quartile when at least half are non-spanning or under defined conditions for cases with broad somatic repeat length distributions. Additional non-spanning reads may be assigned as supporting evidence without contributing to length estimation. Interquartile ranges describe read-level variation.

Allele motif summaries use median per-read counts with cyclic rotations normalized; unexpected motifs are only reported if they are the most common or pass additional consistency and strand support checks. 5mC and 5hmC modifications are extracted from BAM tags for CpGs present in repeat regions and summarized per allele. Outputs include allele-level VCF calls with read support, quality flags and assignment status, per-read TSV results and a tagged BAM file. Repeat-length distributions and read-level visualizations link motif organization and interruptions to allele assignment, read strand and CpG modification, enabling inspection of both the calls and their supporting evidence.

### Locus catalog and adaptive sampling panel

Tandem repeat loci were drawn from STRchive (Hiatt et al. 2025). Each participating site designed its own adaptive sampling target file, reflecting differences in the scope of the testing performed at that center. The Oxford Nanopore panel targeted 104 genes across 28.9 Mb (0.93% of the genome), the University of Washington (UW) panel targeted 201 genes across 55.3 Mb (1.78%), and the Victorian Clinical Genetics Services (VCGS) panel targeted 613 genes across 56.8 Mb (1.83%). The UW and VCGS panels additionally include genes associated with conditions other than repeat expansion disorders, so panel size does not correspond to the number of tandem repeat loci interrogated. In addition, some samples sequenced at VCGS were performed on an ad-hoc basis, and thus did not utilize the same target list as others. Padding around each target also differed between sites, with the UW panel using approximately 100 kb of non-strand-specific padding and the VCGS panel using narrower intervals, so the region targeted at a shared gene is not identical between sites. Seventy-three genes were targeted by all three panels, and the intersection of their coordinates defines a common region of 11.4 Mb, or 0.37% of the genome, which contains the tandem repeat loci used for all cross-site comparisons **(Supplemental Table 1)**. Repeat motifs and pathogenic thresholds used for classification are listed in **Supplemental Table 2**. Unless otherwise stated, results compared across sites are restricted to this shared set.

### Sequencing of Genome in a Bottle Samples

High molecular weight DNA was extracted from HG002–HG007 using the Puregene Cell Kit (Qiagen). For all samples, DNA was quantified using a Qubit fluorometer and fragment size was evaluated using an Agilent Femto Pulse. For WGS, samples were sheared using the Megaruptor 3 (Diagenode) at speed 30 to a fragment length of 25–30 kb. Libraries were prepared using the SQK-LSK114 kit (ONT) and sequenced on PromethION FLO-PRO114M flow cells to at least 93 Gb per sample. For AS, samples were sheared using a VIAFLO 300 pipette (Integra), and libraries were prepared using the SQK-NBD114.24 kit (ONT). Samples were pooled and sequenced on a PromethION 24 using FLO-PRO114P (PromethION Plus) flow cells, with one flow cell used for every three samples in the pool. AS was performed against GRCh38 with the target regions described above, and each pool was sequenced for approximately 72 hours without washing or reloading. Basecalling was performed using Dorado v2.0.0 with the HAC v6.0.0 model. Reads were aligned to GRCh38 using minimap2 (v2.28) (Li 2018) with the map-ont preset, variants were called and phased using Clair3 (v1.0.8) (Zheng et al. 2022) with the ONT R10.4.1 model and the --enable_phasing option, aligned BAM files were haplotagged using LongPhase (v1.7.3) (Lin et al. 2022), and BAM manipulation was performed with SAMtools (v1.19) (Danecek et al. 2021) **(Supplemental Table 3)**. Tandem repeats were genotyped using Mahi-mahi (v0.7.1) with default settings and a locus bed that included *NOTCH2NLC* that was filtered out in downstream analysis.

### Orthogonal genotyping of Genome in a Bottle samples with PacBio HiFi data and TRGT

HiFi reads aligned to GRCh38 for the GIAB samples were obtained from the National Institute of Standards and Technology reference material repository (https://ftp-trace.ncbi.nlm.nih.gov/ReferenceSamples/giab/data/). Revio data were used for the Ashkenazi trio (HG002–HG004) and Sequel II data (PacBio_CCS_15kb_20kb_chemistry2) for the Han Chinese trio (HG005–HG007). TRGT (v5.1.0-ec66463) was run on each sample using the Mahi-mahi target catalog reformatted for TRGT (Dolzhenko et al. 2024). The copy number of each TRGT allele was estimated by dividing the allele length (AL) by the median motif length for each locus and rounding to the nearest integer. This was compared to the copy number reported in the Mahi-mahi TSV output since, by default, the VCF only reports the pathogenic motif for some loci. For composite TRs, the total Mahi-mahi copy number was used for comparison. The shorter allele from both cohorts was labeled as hp1 and the longer allele was labeled as hp2. In some cases, further comparison was done at the TR component level which used the motif counts per allele (MC) values from the TRGT VCF and the component copy numbers in the Mahi-mahi TSV, or the median counts of expanded motifs (MRCN) estimations from the Mahi-mahi VCF.

### Comparison with assembly-derived genotypes in 1000 Genomes samples

We assessed concordance between repeat lengths called by Mahi-mahi (v0.7.1) and by vamos (v3.0.6) (Ren et al. 2023) in 245 samples from the 1000 Genomes Project sequenced on R10.4.1 flow cells (Gustafson et al. 2024) **(Supplemental Table 4)**. Mahi-mahi calls were generated from reads basecalled with the v6.0 HAC model and aligned to GRCh38. For the same samples, we used publicly available vamos calls derived from haplotype-resolved hifiasm (Cheng et al. 2021) assemblies aligned to GRCh38 (Gibson et al. 2026), which cover the STRchive locus set (Hiatt et al. 2025). Of the 72 tandem repeat loci in the Mahi-mahi catalog, 68 overlapped a locus in this set **(Supplemental Table 5)**, and 57 of these retained at least one allele after the filters described below.

Because the two tools can define the boundaries of a repeat differently and may include different motifs in a call, we harmonized locus definitions before comparison. For each locus, we added a constant offset to the vamos count equal to the difference between the repeat counts each tool reports for the GRCh38 reference allele. At composite loci, we compared the equivalent repeat in each tool: at *HTT*, the leading CAG tract, excluding the terminal CAA-CAG interruption and the adjacent CCG tract; at *ATXN8OS*, CTG units only, excluding CTA units; and at *CNBP*, the primary CAGG tract in four-base units, excluding the adjacent CAGA and CA tracts. Coordinates, motifs, length definitions, offsets, and the rationale for each locus are given in **Supplemental Table 5**.

Alleles were matched within each sample and locus by minimizing the sum of absolute differences in repeat length, independently of haplotype labels. We excluded missing or uninterpretable calls and calls carrying the Mahi-mahi LowDepth filter. We also excluded sample–locus comparisons in which the two vamos haplotypes had identical repeat sequences, since these may reflect assemblies in which the haplotypes were not resolved. Finally, we excluded individual alleles whose vamos sequence exactly matched GRCh38 across the repeat interval, since in these cases a repeat-length change may have been placed outside the vamos interval during alignment. Each remaining allele was counted individually. Alleles were classified as concordant if they differed by no more than one repeat unit; larger differences were expressed as a percentage of the harmonized vamos length and grouped as 10% or less, more than 10% to 25%, or more than 25%. For loci at which any allele differed by more than 10%, we inspected both tools’ allele lengths, the offset applied, Mahi-mahi read visualizations, and alignments of the assemblies and raw reads to GRCh38 with the vamos repeat interval marked.

### Coriell reference lines

DNA for sequencing was obtained either directly from the Coriell Institute or extracted from Coriell cell lines using the Puregene Cell Kit (Qiagen) **(Supplemental Table 6)** and sequenced by whole-genome sequencing (WGS) and adaptive sampling (AS) on PromethION flow cells and AS on PromethION Plus flow cells. For all samples, DNA was quantified using a Qubit fluorometer and fragment size was evaluated using an Agilent Femto Pulse. For AS on PromethION flow cells, samples were prepared with the Multiplex Adaptive Sampling Protocol, which included G-tube shearing to approximately 6–8 kb, and sequenced with three samples per flow cell for 72 hours, with one wash performed after approximately 24 hours of sequencing. For AS on PromethION Plus flow cells, the same sample preparation was performed, with three samples sequenced per flow cell for 100 hours and no washes. For AS of Coriell samples the same bed file was used across flow cells, with the entirety of the genes containing repeat expansions flanked by 20 kb strand-specific padding, targeting 0.93% of the genome. For WGS, samples were sheared in the same way as the AS samples to enable direct comparison, libraries for sequencing were prepared with the Ligation Sequencing Kit (SQK-LSK114, ONT) and run as one sample per flow cell on a PromethION flow cell for 72 hours with no washes. PromethION data were basecalled with MinKNOW (v26.01.21) using the v5.2 HAC model, and dorado (v2.1.2+8b8fc5d) for v5.2 SUP, and v6.0 HAC models, all with ‘5mCG_5hmCG’ methylation detection, and PromethION Plus data with dorado (v2.1.2+8b8fc5d) and the v1.0 HAC PromethION Plus model, followed by analysis with EPI2ME wf-human-variation (v2.8.1, ONT) and Mahi-mahi (v0.7.1, ONT). Orthogonal genotypes for all 14 lines were taken from the publicly released PacBio PureTarget repeat expansion panel results, generated with TRGT (Dolzhenko et al. 2024) on both Revio and Vega instruments (https://www.pacb.com/technology/puretarget/). For comparison with TRGT we used the median number of repeat units per allele (REPCN) from the Mahi-mahi VCF. At *FMR1*, which was genotyped in every line, we compared the mean CpG methylation that each tool reports for each allele.

### Sequencing of laboratory samples

DNA from blood samples at the UW site was extracted using the QIAsymphony with the QIAsymphony DSP DNA Midi Kit (Qiagen) **(Supplemental Table 7)**. For all samples DNA was quantified using a Qubit Fluorometer and fragment size was evaluated using an Agilent Femto Pulse. Samples were sheared using the Megaruptor3 (Diagenode) at speed 31. Libraries were prepared using the SQK-HTB114.96 library kit (ONT). Samples were pooled and sequenced on a PromethION 24 using FLO-PRO114P (PromethION Plus) flow cells, with one flow cell used for every three samples in the pool. AS was performed using the GRCh38 reference genome with target regions as described above. Each sample was sequenced for approximately 72 hours without washing or reloading. Basecalling was performed using Dorado v2.0.0 with the HAC v6.0.0 model. Reads were aligned to the GRCh38 reference genome using minimap2 (v2.28) with the map-ont preset. Variant calling and phasing was performed using Clair3 (v1.0.8) with the ONT R10.4.1 model and --enable_phasing option. Aligned BAM files were haplotagged using LongPhase (v1.7.3). BAM file manipulation was performed using SAMtools (v1.19). Tandem repeats were genotyped using Mahi-mahi (v0.7.1) with default settings and a locus bed that included *NOTCH2NLC* that was filtered out in downstream analysis. At UW, *ATXN1* interruption status had been determined during orthogonal testing by PCR followed by SfaNI restriction digest, which cuts at the GCATC sequence created by a CAT interruption within the CAG tract.

DNA for samples sequenced by University Medical Center Utrecht was extracted from peripheral EDTA blood using chemagic (Revvity) isolation **(Supplemental Table 8)**. DNA was fragmented using FastPrep homogenization (MP Biomedicals). Library prep was performed using standard protocol for WGS (SQK-LSK114-XL) on the Callisto sample prep system (Volta labs). Each sample was sequenced on a PromethION 24 using two R10.4.1 flowcells (FLO-PRO114M). Sequencing was performed for 72 hours without washing, targeting >30X genome coverage per sample combining the two flowcells. Basecalling (using the dna_r10.4.1_e8.2_400bps_HAC@v5.2.0 model with 5hmC & 5mC in all contexts) and mapping (GRCh38) were performed during sequencing using MinKNOW software (v25.09.16). Data from both flowcells were combined for secondary analysis using the EPI2ME wf-human-variation (v2.7.2) workflow (https://github.com/epi2me-labs/wf-human-variation/), followed by STR calling using Mahi-mahi (v0.7.1) with default settings.

DNA for samples sequenced by VCGS were a combination of archival clinical samples and research positive control samples, extracted from peripheral blood collected in EDTA tubes using either QIASymphony automated DNA extraction (Qiagen) or Mag-Bind DNA Blood and Tissue kit (Omega Bio-tek) **(Supplemental Table 9)**. Concentration was measured using the Qubit dsDNA Broad Range assay (Thermo Fisher Scientific) and integrity was assessed using the Agilent Genomic DNA ScreenTape assay on the TapeStation platform. Samples with at least 1 µg of DNA and a DNA Integrity Number above 7.0 were considered suitable for long-read library preparation, and DNA was normalized to 1 µg in 50 µl (20 ng/µl). To optimize read length for AS, DNA was mechanically sheared to approximately 10–15 kb using a Geno/Grinder 2010 (Cole-Parmer) at 1600 rpm for 5 minutes, after which samples proceeded directly to library preparation. Sequencing libraries were prepared using the SQK-NBD114.24/96 kit with a modified high-throughput barcoding method. Libraries were normalized by pooling 2 µl of each library and sequencing the pool on a low-pore-count PromethION flow cell (<5,000 pores) for 8–12 hours; passed bases per sample were used to determine barcode normalization for the main run. Libraries were then pooled at four samples per PromethION flow cell (FLO-PRO114M) and sequenced for 72 hours with one to two washes as required, targeting approximately 30x coverage per sample across enriched regions. Data were processed with an in-house validated pipeline based on the ONT human variation workflow with basecalling performed on instrument (MinKNOW v25.09), with v4.3 or v5.2 models and 5mCG_5hmCG methylation calling as indicated per sample, followed by alignment to GRCh38 using minimap2 (v2.24), variant calling with Clair3 (v1.0.7), phasing using LongPhase (v1.7), and repeat expansion calling with Mahi-mahi (v0.7.1). The workflow can be found in https://github.com/ssadedin/warpy.

### Orthogonal comparison and statistics

Orthogonal genotypes were classified as one of four types: exact values, lower bounds (reported as greater than a threshold), upper bounds (reported as less than a threshold), or ranges. Only exact values were used for quantitative concordance, and alleles were sorted by size before comparison. Genotypes for which Mahi-mahi reported a single allele where the orthogonal assay reported two were excluded from allele-level comparisons. Ninety-five percent confidence intervals for proportions were calculated with the Wilson score method. Because the precision of PCR-based assays varies with allele size, concordance was computed separately for alleles below and above 100 repeat units. Category-level concordance (normal, intermediate or premutation, pathogenic) was computed against the locus-specific thresholds in **Supplemental Table 2** for every genotype: exact results were concordant when both calls fell in the same category, lower-bound results when the Mahi-mahi call fell in the same or a more severe category, and upper-bound results when it fell in the same or a less severe category. Technical reproducibility was assessed in sample-locus combinations sequenced as two or three independent libraries, restricting the primary analysis to series in which every library was basecalled with the same model. For replicates where a sample was sequenced as more than one library, the libraries were collapsed to a single genotype for all concordance analyses by taking the per-allele median across libraries basecalled with the most recent model, so that each sample–locus combination contributed once; individual libraries were used only for the analysis of technical reproducibility. At *PABPN1*, the referring laboratory reported the number of GCG triplets, whereas Mahi-mahi reports the full GCN tract; orthogonal results were converted to GCN units (GCG count plus four) before comparison.

## RESULTS

### Mahi-mahi workflow and target catalog

Mahi-mahi genotypes tandem repeats from aligned ONT reads using a curated locus catalog. For each target, reads spanning the repeat and its flanking sequence are extracted, the repeat structure of every read is decomposed into its constituent motifs, and reads are clustered into alleles. For each allele, Mahi-mahi reports repeat length, motif composition, and the position and identity of interruptions, together with read-level flags that identify reads carrying systematic, motif-biased basecalling error. The output of Mahi-mahi is a per-sample VCF, a per-read tabular summary of every genotyped locus, and static visualizations of allele clustering and repeat structure and CpG modifications **(Supplemental Figure 1)**. We evaluated the performance of Mahi-mahi across four cohorts and two experimental configurations: WGS and AS applied to six GIAB reference samples, 245 samples from the 1000 Genomes Project, 14 Coriell reference lines carrying known expansions, and 103 samples from three laboratories (**Table 1**).

**Table 1.**
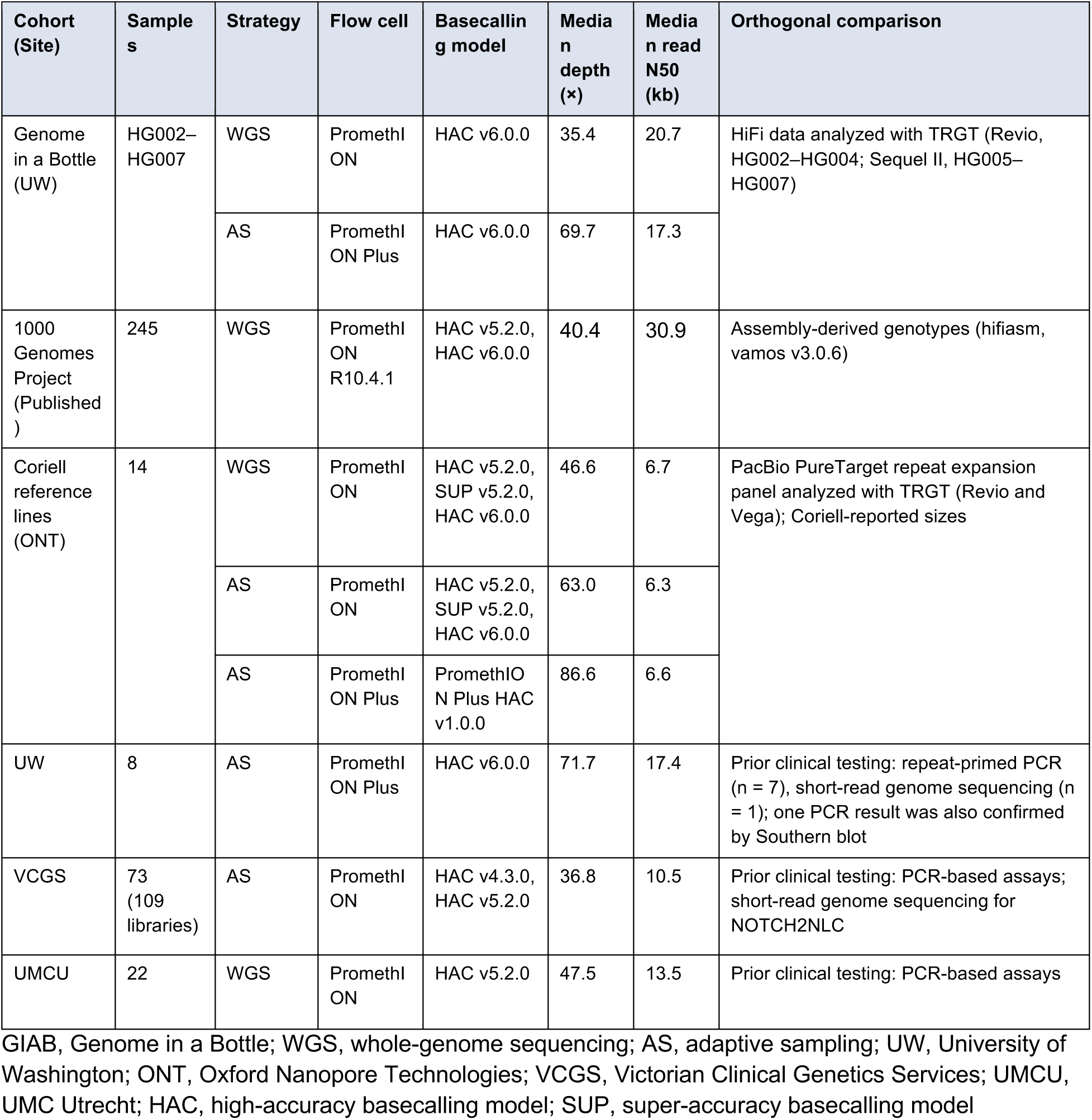
Cohorts used to evaluate Mahi-mahi. Depth is genome-wide for whole-genome sequencing and on-target for adaptive sampling. At VCGS, 20 samples were sequenced as two or three libraries.

### Concordance of Mahi-mahi between whole-genome sequencing and adaptive sampling

We first tested whether AS (targeted enrichment) degrades genotyping relative to WGS. Six GIAB samples (HG002–HG007) were sequenced by both approaches. WGS achieved a median depth of 35.4x (range 31.0–41.2x) with a median read N50 of 20.7 kb, while AS, running three samples per flowcell, achieved a median on-target depth of 69.7x (range 46.0–81.2x) against a median off-target depth of 2.6x, representing approximately 27-fold enrichment (range 24–31-fold) with a median read N50 of 17.3 kb (**Figure 1A, Supplemental Table 3)**. Across 72 catalog loci, WGS and AS calls were identical for 821 of 832 alleles and within one repeat unit for 829 (99.6%; 95% CI 98.9–99.9%); the three larger differences were at *CNBP* in HG003 (50 repeat units by WGS versus 47 by AS), at *BEAN1* in HG004 (526 versus 528), and at the expanded *RFC1* allele in HG007 (403 versus 398) **(Supplemental Table 10)**.

**Figure 1.**
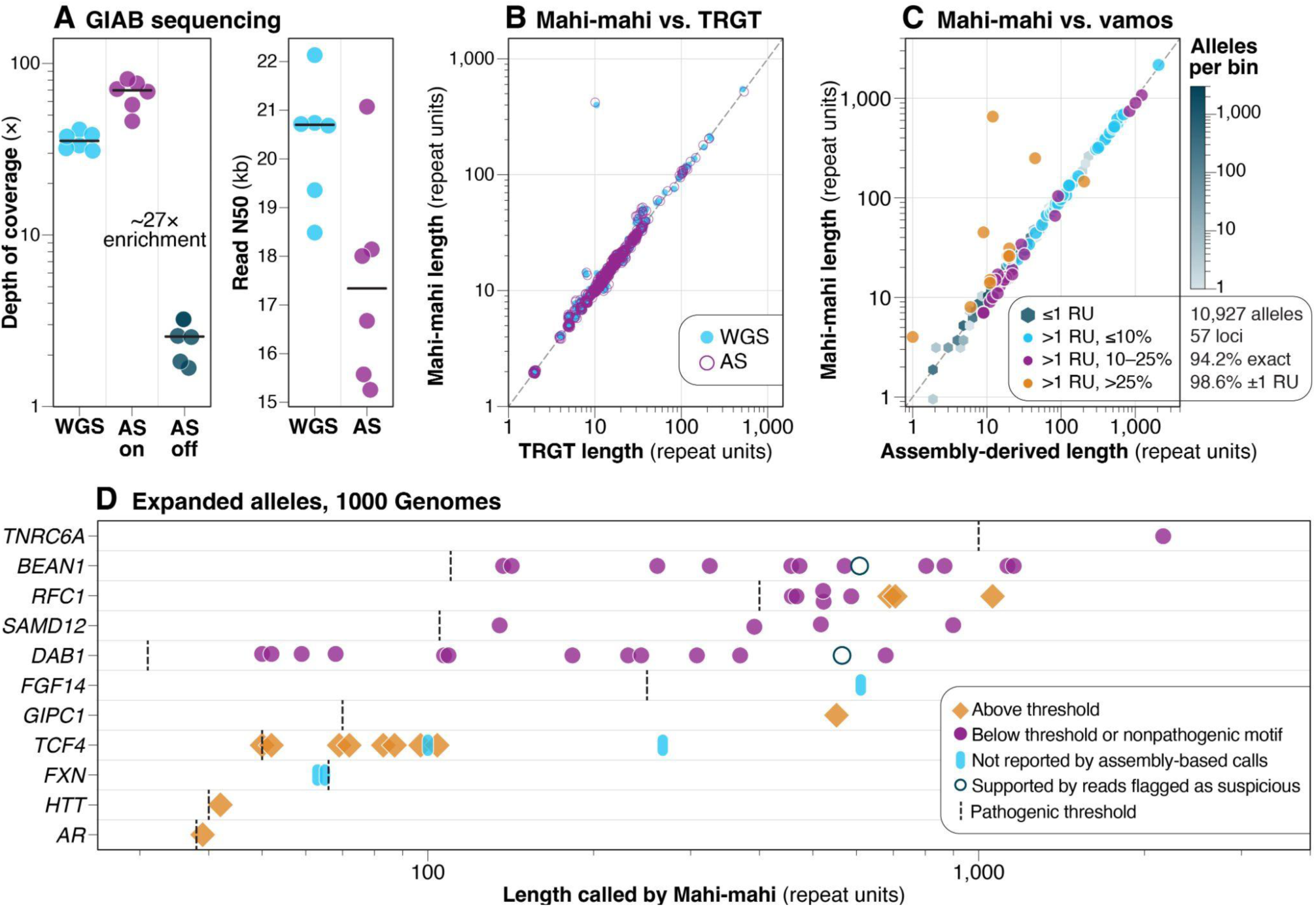
Benchmarking Mahi-mahi in Genome in a Bottle and 1000 Genomes samples. **(A)** Depth and read N50 for whole-genome sequencing (WGS) and adaptive sampling (AS) in HG002–HG007, with AS depth shown separately on and off target. Horizontal lines mark the median. **(B)** Mahi-mahi repeat length (repeat units) from ONT WGS and AS versus TRGT repeat length from HiFi data (Revio for HG002–HG004, Sequel II for HG005–HG007). **(C)** Mahi-mahi repeat length versus vamos repeat length from haplotype-resolved assemblies in 245 samples from the 1000 Genomes Project. **(D)** Expanded alleles called by Mahi-mahi in the 1000 Genomes cohort, by locus, at the length Mahi-mahi reports. These comprise the 49 alleles previously reported from assemblies and five not reported there (cyan; two at *TCF4*, two at FXN, one at FGF14; **Supplemental Figure 7**). Ticks mark the pathogenic threshold **(Supplemental Table 2)**; at *RFC1* this is the AAGGG threshold of 400 repeat units. Orange points are at or above the threshold. Purple points are below it or carry a nonpathogenic motif: the reference motif at *BEAN1*, *DAB1*, *SAMD12* and *TNRC6A*, or AAAGG at *RFC1*, which is pathogenic only at 600 repeat units or more. One of the three orange diamonds at RFC1 marks the only AAAGG allele above 600 (HG02220, 706 repeat units). All *RFC1* alleles shown are heterozygous. Allele lengths are given in **Supplemental Table 12**.

### Comparison with TRGT in Genome in a Bottle samples

We next compared Mahi-mahi genotypes with TRGT genotypes generated from HiFi data for the same samples. Of the 832 TRGT alleles across 72 loci, Mahi-mahi reported a genotype for 643; the remaining 189 were unavailable for comparison, 186 because both TRGT alleles were the same size and by design Mahi-mahi reported a single allele, and three (*ATXN8OS* in HG003, *FXN* in HG004, and *TCF4* in HG006) because Mahi-mahi reported a single allele where TRGT reported two alleles one repeat unit apart. Without adjustment, Mahi-mahi calls from WGS data matched TRGT exactly for 477 of 643 alleles (74.2%) and were within one repeat unit for 587 (91.3%), with every allele within one repeat unit at 63 of 72 loci (**Figure 1B, Supplemental Figure 2, Supplemental Table 10)**. Excluding *CNBP*, *FGF14*, and *GOLGA8A,* at which the two tools define the repeat differently (below), 582 of 609 alleles (95.6%; 95% CI 93.6–96.9%) were within one repeat unit. Most one-unit differences were constant within a locus: −1 at HTT and +1 at *EIF4A3*, *FAM193B*, *MARCHF6*, *RAPGEF2*, *RILPL1*, *TNRC6A*, *TYMS*, and *YEATS2*. These account for 94 of the 110 one-unit differences and point to differences in boundary definition rather than sizing error.

Three loci were consistently discordant by more than one repeat unit: *CNBP, FGF14,* and *GOLGA8A*. *CNBP* and *GOLGA8A* are composite loci. Comparing the median counts of expanded motifs (MRCN) value from the Mahi-mahi output for the primary motif for *CNBP, FGF14,* and *GOLGA8A* to the TRGT motif counts per allele (MC) value for the same motif improved agreement at all three **(Supplemental Figure 3)**, indicating that the differences arise from how interruptions or the boundaries between components are handled. Similarly, at *HTT* the boundary between the CAG tract and the adjacent CAACAG and CCG units was placed differently, so that Mahi-mahi calls were one repeat unit shorter than the TRGT estimate in every allele, and at *RAI1* and *SAMD12* Mahi-mahi calls were a constant 7 bp longer, which rounds to two repeat units of these pentanucleotide motifs and accounts for 20 of the 27 remaining alleles outside one repeat unit.

Comparing motif counts, rather than copy numbers estimated from allele length, also revealed alleles composed largely of a motif absent from the catalog entry for the locus, at *BEAN1* in HG004 and at *STARD7* and *YEATS2* in several samples. TRGT recovered the length of these alleles in base pairs, and in some cases their consensus sequence, but returned a reduced or zero copy number because it counts only the motifs listed in the catalog; the 2,588-bp *BEAN1* allele in HG004, for example, received a TRGT motif count of zero, and a more recent TRGT release did not change this. Mahi-mahi total copy numbers at all three loci tracked allele length because they sum every repeat unit in the decomposed reads. At *BEAN1*, Mahi-mahi also identified the uncatalogued motif, reporting the long HG004 allele as approximately 520 TACAA units, a nonpathogenic motif distinct from the TGGAA insertion that causes SCA31 (spinocerebellar ataxia type 31, OMIM #117210) **(Supplemental Figure 4)**. At *STARD7* and *YEATS2*, Mahi-mahi sized the alleles but did not assign the uncatalogued units to a motif. Because neither catalog included these motifs, we report these alleles separately rather than as discordances.

Additional differences attributed to sequencing include *RFC1* in HG007 and *ZNF713* in HG005, HG006 and HG007. At the *RFC1* locus Mahi-mahi called an expanded allele of approximately 400 repeat units in HG007 that TRGT did not report; on review of the PacBio data we found no reads on that haplotype that extended into the expansion, consistent with undersampling of the long allele in the Sequel II data **(Supplemental Figure 5)**. At the longer *RFC1* allele in HG004, the copy number estimate was 85 units from TRGT and 74–75 units from ONT AS and WGS data. At *ZNF713*, nine of twelve alleles matched TRGT exactly, but in the remaining three individuals (HG005, HG006, and HG007), TRGT reported both alleles as equal in copy number (8 and 8, 11 and 11, 8 and 8) while Mahi-mahi called the longer allele at 14 units in each. These are also the only *ZNF713* alleles whose TRGT allele length range (ALLR) does not overlap the Mahi-mahi AS range, by 5 to 13 bp **(Supplemental Figure 3)**.

### Concordance with assembly-derived genotypes in 1000 Genomes samples

We applied Mahi-mahi to 245 samples from the 1000 Genomes Project and compared the resulting allele length distributions with assembly-derived genotypes generated from haplotype-resolved hifiasm assemblies with vamos over the same locus set. Across 10,927 retained allele comparisons at 57 loci in 245 samples, Mahi-mahi and assembly-derived vamos calls matched exactly for 94.2% of alleles and to within one repeat unit for 98.6% **(10,776 of 10,927; 95% CI 98.4–98.8%; Figure 1C, Supplemental Table 11)**. Among 97 alleles differing by more than 10%, manual review identified systematic differences at *ATN1* (longer Mahi-mahi estimates associated with interruptions), *CNBP* (complex repeat structure affecting assembly-based calls) and *LRP12* (deletions extending outside vamos intervals). Long alleles, including at *RFC1*, also differed because Mahi-mahi estimates were near the median repeat length across reads, whereas assembly-derived lengths were nearer the upper end of the distribution. Excluding *ATN1*, *CNBP* and *LRP12* increased agreement to 99.3%. The three largest relative discordances, two at *RFC1* and one at *YEATS2*, were each an expanded allele called by Mahi-mahi and supported by both the raw reads and one assembly haplotype but absent from the vamos call: in HG04211 (*RFC1*) and HG04033 (*YEATS2*), the inserted sequence was placed just outside the vamos interval during alignment, and in HG00253 (*RFC1*) the reason the expansion was not called could not be determined **(Supplemental Figure 6)**.

Mahi-mahi recovered all 49 expanded alleles previously reported in this cohort across nine loci (*AR*, *BEAN1*, *DAB1*, *GIPC1*, *HTT*, *RFC1*, *SAMD12*, *TCF4*, and *TNRC6A*) (Gibson et al. 2026), with a median difference of 1.9% from the assembly-derived length and 44 within 10% **(Supplemental Table 12)**; a fiftieth previously reported expansion, a second *BEAN1* allele in HG00109, was not supported by the reads and reflected an assembly error. Most of these alleles are long but composed of the reference motif or another motif not associated with disease, and are therefore not expected to be pathogenic despite their length. The largest alleles recovered exceeded 1,000 repeat units at *BEAN1*, *RFC1*, and *TNRC6A*. Among the eight individuals carrying *RFC1* alleles above 400 repeats, all of which were heterozygous, two carried the AAGGG motif that causes CANVAS (cerebellar ataxia, neuropathy, and vestibular areflexia syndrome, OMIM #614575) (Cortese et al. 2019) and six carried AAAGG alleles of comparable or greater size (**Figure 1D**). AAAGG expansions are usually benign, but alleles of 600 or more repeat units are considered pathogenic (Hiatt et al. 2025), and one allele here (HG02220, 706 repeat units) exceeds that size. Because CANVAS is recessive, none of these heterozygous alleles is expected to cause disease on its own, but the *RFC1* calls show that both motif and length are needed to interpret an allele. At *DAB1*, assembly-derived calls for two individuals reported GAAAT units, which on the opposite strand are the ATTTC insertion that causes SCA37 (spinocerebellar ataxia type 37, OMIM #615945) (Seixas et al. 2017). The assembly sequence in fact contained GGAAT, the product of a strand-specific basecalling error, and vamos reported it as GAAAT because, of the two motifs it considers at this locus (AAAAT and GAAAT), GAAAT was the closer match. Because the miscall recurred in every read from the affected strand, assembly reinforced it rather than removing it. A systematic basecalling error combined with a genotyper restricted to a small set of motifs can therefore produce sequence that mimics a pathogenic allele. Mahi-mahi instead masked reads whose motif composition was supported by only one strand and called the canonical AAAAT motif in both individuals (**Figure 5F**).

Mahi-mahi also identified expanded alleles in five individuals that were not reported in the assembly-based analysis **(Supplemental Figure 7)**. Two, at *TCF4* in HG00231 and HG02351, were absent from the assembly-based calls because the inserted sequence was placed just upstream of the vamos interval during alignment and, for the same reason, were removed from concordance analysis above by the filter for reference-matching sequence. Two others, at *FXN* in HG03757 and HG04235, are composite alleles of 65 and 63 repeat units in which tracts of GAA triplets are separated by a similar number of GGA triplets. Expanded GAA–GGA composite alleles were recently shown to be common among pathogenic *FXN* alleles in Friedreich ataxia (OMIM #229300) and to be missed by conventional PCR-based testing (Devore et al. 2026); the alleles observed here are much shorter than those described, their longest pure GAA tracts fall below the pathogenic threshold, and their significance is uncertain. The fifth, at *FGF14* in HG00556, is an allele of 611 repeat units interrupted throughout by GCA units. Because pathogenic SCA27B (spinocerebellar ataxia type 27B, OMIM #620174) alleles are uninterrupted GAA tracts and non-pathogenic alleles of comparable length are typically interrupted (Pellerin et al. 2023), this allele is unlikely to be pathogenic, although to our knowledge interruptions of this composition have not been described.

### Evaluating pathogenic expansions in Coriell reference lines

We assessed performance on pathogenic alleles using 14 Coriell reference lines with publicly released PacBio PureTarget genotypes generated with TRGT on both the PacBio Revio and Vega platforms. These 14 lines span six loci (*HTT*, *ATXN1*, *DMPK*, *PABPN1*, *RFC1*, and *FXN*) and include expanded alleles from 13 to more than 1,000 repeat units (**Figure 2E, Supplemental Table 6)**. Each line was sequenced three ways with nanopore sequencing: WGS and AS on PromethION flow cells, and AS on PromethION Plus flow cells (**Table 1**, **Figure 2A**).

**Figure 2.**
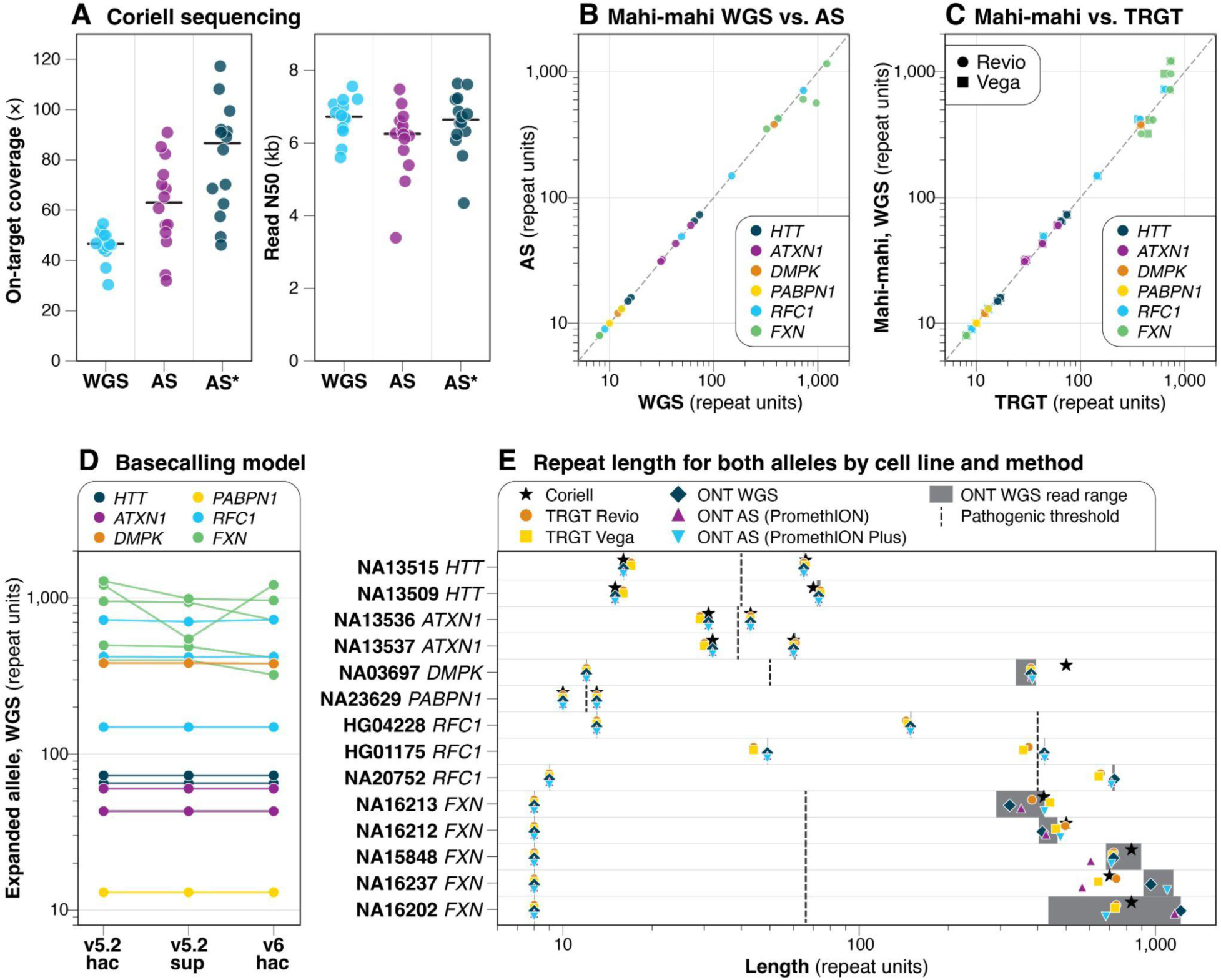
Evaluation of Mahi-mahi in Coriell reference lines. **(A)** On-target depth and read N50 for three ONT configurations: whole-genome sequencing (WGS) and adaptive sampling (AS) on PromethION flow cells, both basecalled with v6.0 HAC; and adaptive sampling on PromethION Plus flow cells (AS*), basecalled with the PromethION Plus v1 HAC model. **(B)** Mahi-mahi repeat length from WGS versus AS (PromethION), colored by locus. **(C)** Mahi-mahi repeat length from WGS versus TRGT motif count from PacBio PureTarget data on Revio (circles) and Vega (squares). **(D)** Mahi-mahi expanded allele length under the v5.2 HAC, v5.2 SUP and v6.0 HAC models; lines connect calls from the same cell line. **(E)** Both alleles for each cell line by method, with Coriell-reported sizes where available. Coriell reports only the expanded allele for the *DMPK* and *FXN* lines and no size for the three *RFC1* lines (HG01175, HG04228, NA20752). Shaded bars show the range of repeat lengths across reads assigned to each allele in the WGS data; at many alleles they are hidden by the markers. Colors in C–E follow B. *PABPN1* is shown in GCN units, so the Coriell genotype of 6 and 9 GCG repeats is plotted as 10 and 13.

Mahi-mahi identified the expected expansion in all 14 lines under all three sequencing configurations and basecalling models. At *HTT*, *ATXN1*, *DMPK*, and *PABPN1*, every allele called by Mahi-mahi from whole-genome data was within two repeat units of the TRGT calls, and WGS and AS calls for both PromethION and PromethION Plus flow cells were identical or nearly identical at all 12 alleles at these loci (**Figure 2B, 2C, Supplemental Table 6)**. For those lines in which Coriell reports an exact genotype, Mahi-mahi was identical at 8 of 10 alleles and was within one repeat unit at 9 of 10, compared with 3 of 10 and 7 of 10 for TRGT. The differences were concentrated at shorter alleles, where Mahi-mahi returned the Coriell-reported genotype exactly at both *ATXN1* lines (32 and 31 repeats, against TRGT calls of 30 and 29) and at both *HTT* lines (16 and 15 repeats, against TRGT calls of 17 and 16). The single locus at which both long-read platforms disagreed with Coriell was the expanded *HTT* allele in NA13509, called at 73 repeats by Mahi-mahi and 74 by TRGT against a reported 70, which suggests that the reference value is not accurate rather than an error in both callers.

At *RFC1*, Mahi-mahi called the expanded allele 3% to 13% larger than the TRGT motif count in all three lines (421 versus 373 repeats in HG01175, 149 versus 144 in HG04228 (**Figure 5D**), and 727 versus 653 in NA20752). The TRGT allele length in base pairs, however, corresponds to 406, 149, and 681 repeat units, within 7% of the Mahi-mahi call in every case. As at the GIAB loci described above, the discrepancy therefore likely reflects the portion of the *RFC1* expansion composed of motifs other than those counted by TRGT, rather than a difference in the length of the allele.

Nanopore reads are converted from raw electrical signal to sequence by a basecaller using machine learning models. ONT periodically releases new basecalling models, each available at several accuracy tiers (for example, high-accuracy, HAC, and super-accuracy, SUP). Because the raw signal is retained, the same data can be re-basecalled with a newer model, so the model used can differ between laboratories, between samples sequenced at different times, and between analyses of the same sample. To test whether this affects Mahi-mahi calls, we basecalled the Coriell whole-genome and adaptive sampling data with three models (v5.2 HAC, v5.2 SUP, and v6.0 HAC) and compared the resulting genotypes. Basecalling model had little effect except at the *FXN* locus. At the other five loci, the v5.2 HAC, v5.2 SUP, and v6.0 HAC models returned identical calls for 29 of 36 alleles, with the largest difference being 21 repeat units (3%) between the v5.2 HAC and v6.0 HAC calls at the expanded *RFC1* allele in NA20752 (**Figure 2D**). Across the five *FXN* cell lines, calls for the expanded allele varied substantially between basecalling models, sequencing configurations, and platforms; in NA16202, for example, Mahi-mahi called the expansion at 547 to 1,217 repeats depending on the model, TRGT at 731 to 738, while Coriell reports approximately 830 (**Figure 2E**). When the whole-genome data basecalled with the v5.2 SUP model, which v0.7.1 called at 547 repeats, were reanalyzed with a newer version of Mahi-mahi (v1.0) that accommodates a 39-bp deletion in the downstream flank of this allele, the call was 816 repeat units, close to the Coriell-reported size **(Supplemental Figure 8)**. This variation was mirrored within samples: the repeat lengths of individual reads assigned to a single expanded *FXN* allele spanned up to 782 repeat units in one line, and TRGT likewise reported base-length ranges spanning more than 1,000 bp at the same alleles. Such broad within-sample distributions are consistent with somatic instability of the GAA expansion in lymphoblastoid cell lines (Long et al. 2017), and indicate that for these alleles the expansion itself is reproducibly detected while its size is not a single stable quantity. Every method classified every *FXN* expansion as pathogenic.

Because every line was also genotyped at *FMR1*, these data allowed allele-resolved methylation to be assessed using X-chromosome inactivation as an internal control. In all seven XY lines, Mahi-mahi reported a single unmethylated *FMR1* allele (mean methylation 0.02 or less), as did TRGT. In the two XX lines homozygous for a 30-repeat allele, Mahi-mahi reported intermediate methylation (0.46 and 0.53), consistent with a mixture of active and inactive X chromosomes and with TRGT (0.45 and 0.48). In four of the five heterozygous XX lines, the two tools agreed on which allele was more highly methylated, although Mahi-mahi estimated higher methylation than TRGT for the less-methylated allele in three of these (HG01175, NA16237, and NA23629); in NA16212, Mahi-mahi reported distinct methylation of the two alleles (0.13 and 0.76 in whole-genome data; 0.25 and 0.84 with adaptive sampling) where TRGT reported similar methylation of both (0.46 and 0.48 on Revio; 0.40 and 0.57 on Vega) **(Supplemental Figure 9)**. *FMR1* allele lengths from ONT WGS and AS data agreed in every line for which both were available, and per-allele methylation estimates differed by no more than 0.12. Mahi-mahi *FMR1* lengths exceeded the TRGT CGG count by one or two repeat units in 54 of 56 allele comparisons, consistent with Mahi-mahi counting the AGG interruptions within the repeat, which the TRGT CGG count excludes.

### Performance against orthogonal testing

We evaluated Mahi-mahi against prior results using 103 samples from three laboratories: 8 samples sequenced by adaptive sampling at UW, 73 samples (109 libraries) sequenced by adaptive sampling at the VCGS, and 22 samples sequenced by whole-genome sequencing at UMC Utrecht **(Supplemental Tables 7–9)**. Twenty VCGS samples were sequenced as two or three independent libraries to evaluate replicability, and for all concordance analyses these were collapsed to a single genotype per sample and locus so that each sample contributed once. Together these yielded 180 sample–locus genotypes across 23 genes, with median on-target depths of 71.7x, 36.8x, and 47.5x and median read N50 of 17.4, 10.5, and 13.5 kb at UW, VCGS, and UMCU, respectively (**Figure 3A, 3B)**.

**Figure 3.**
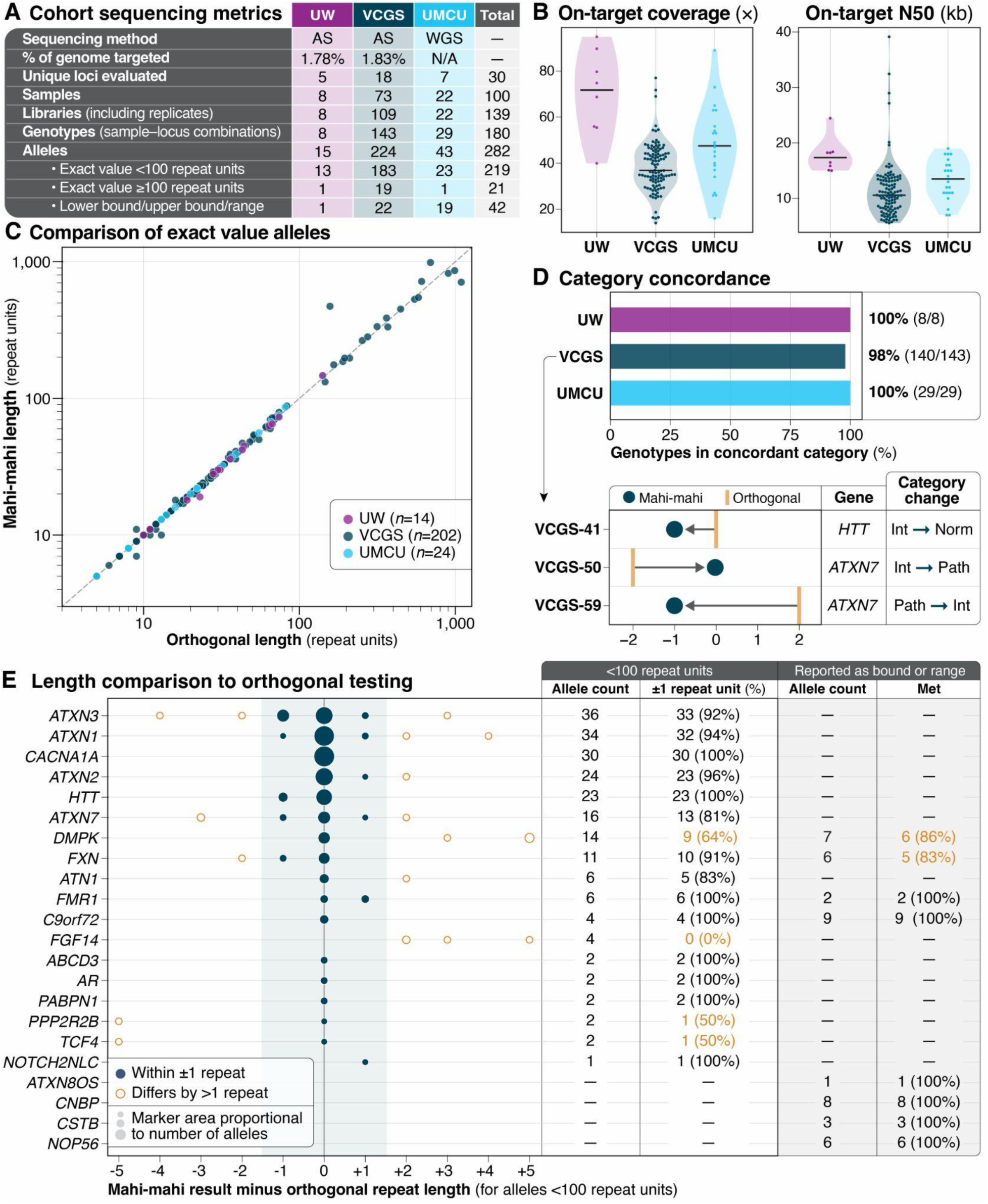
Sample cohort performance. **(A)** Cohort summary by site including alleles with exact values or defined bounds/range. A genotype is one sample–locus combination after replicate libraries are collapsed. An additional 73 alleles were not scored in Figure 3 because orthogonal and Mahi-mahi calls could not be unambiguously matched or the orthogonal repeat length was approximate rather than exact or a defined bound. **(B)** On-target depth and read N50 by site, with one point per library. Horizontal lines mark the median. **(C)** Mahi-mahi versus orthogonal repeat length for alleles from genotypes in which every orthogonal value was exact and Mahi-mahi reported the same number of alleles (*n* = 240 alleles), plotted on logarithmic axes with the line of identity dashed. The 24 genotypes in which Mahi-mahi reported a single allele are not plotted. **(D)** Concordance of normal (norm), intermediate (int), and pathogenic (path) categories by site (*n* = 180 genotypes), including genotypes reported as lower bounds, upper bounds, or ranges. A lower-bound result was scored concordant when the Mahi-mahi call fell in the same or a more severe category, and an upper-bound result when it fell in the same or a less severe category. Below, each discordant genotype is plotted relative to the category threshold it crosses, with the orthogonal result shown as a line and the Mahi-mahi call as a circle. VCGS-59 gave the same result from three independent libraries. **(E)** Agreement with orthogonal testing by locus for the 219 alleles below 100 repeat units in panel C. Each point is the difference between the Mahi-mahi call and the orthogonal result for one allele, in repeat units. The bounds met column counts alleles reported orthogonally only as a lower bound, upper bound, or range (*n* = 42 alleles, 40 met), and shows how many the Mahi-mahi call agreed with. A call agrees with a bound if it is at or above a lower bound, at or below an upper bound, or within a range. For example, the *FXN* genotype of VCGS-46 was reported orthogonally as “>85, >85” and called by Mahi-mahi at 827 and 429 repeat units, so it adds two met bounds to that column and no points to the plot. A value in the ±1 column is orange when fewer than 80% of that locus’s alleles fall within one repeat unit, and a value in the bounds met column is orange when any bound was not met. *PABPN1* orthogonal results were converted from GCG to GCN units (GCG count plus four) before comparison.

Mahi-mahi detected an expanded allele in all 71 sample-locus combinations with a known pathogenic expansion, and called 70 of these in the pathogenic range; the exception, *ATXN7* in VCGS-59, is described below. Agreement on allele size depended on the precision of the comparison assay. Among alleles below 100 repeat units with an exact orthogonal value (n = 219), Mahi-mahi agreed exactly for 76.3% (95% CI 70.2–81.4%) and to within one repeat unit for 90.0% (95% CI 85.3–93.3%) (**Figure 3C**). Because PCR-based assays typically report sizes to within one repeat in this range, this level of agreement is at the resolution limit of the comparison assay. For alleles of 100 repeat units or more (n = 21), the median difference was 6.2%, consistent with the wider tolerance of the assay at these sizes. Of 42 alleles reported only as a lower bound, an upper bound, or a range, the Mahi-mahi call agreed with 40; the exceptions were the expanded *DMPK* allele in VCGS-04, called at 1,072 repeats against a reported size of more than 1,100, and the expanded *FXN* allele in VCGS-12, called at 705 repeats against a reported range of 730–770. In both cases the two values are well above the pathogenic threshold, and both methods classified the allele as pathogenic (**Figure 3E**).

We also assessed agreement at the level of the category a laboratory would report to identify any differences between category assignment and the Mahi-mahi result. Mahi-mahi assigned the same category as the referring laboratory for 177 of 180 genotypes (98.3%; 95% CI 95.2–99.4%): 8 of 8 at UW, 140 of 143 at VCGS, and 29 of 29 at UMCU (**Figure 3D**). In each discordant case the two calls differed by three repeat units or fewer and fell on either side of a category threshold. An *HTT* allele reported at 27 repeats was called at 26 in all three libraries from VCGS-41, placing it just below the intermediate range; an *ATXN7* allele reported at 35 repeats was called at 37 in VCGS-50 (only one library was sequenced from this individual), placing it at the pathogenic threshold; and an *ATXN7* allele reported at 39 repeats was called at 36 by Mahi-mahi in all three libraries from VCGS-59, placing it in the intermediate range.

Disagreements with the orthogonal result were not uniformly distributed across loci (**Figure 3E**). At *CACNA1A*, all 30 alleles below 100 repeats matched the orthogonal result exactly, and at *ATXN2*, 22 of 24 did. In contrast, 10 of 36 *ATXN3* alleles and 5 of 23 *HTT* alleles were called one repeat smaller than the orthogonal result (**Figure 5C, Supplemental Figure 10)**, and all four *FGF14* alleles below 100 repeats were called larger. The offsets that are consistent in direction within a locus point to differences in how the repeat boundary is defined by Mahi-mahi and by the orthogonal assay rather than to sizing error, and the *ATXN7* discordance in VCGS-59, in which all three libraries agreed with one another but not with the orthogonal result, illustrates this: both of its alleles were called three repeat units shorter than reported (10 and 36 versus 13 and 39). In the cases in which orthogonal testing reported two alleles of identical size, Mahi-mahi generally returned a single allele, indicating a homozygous repeat length. Allele clustering failed in one sample: at *ATXN1* in VCGS-53, which was sequenced three times and for which orthogonal testing reported alleles of 29 and 30 repeats, two of three libraries returned a single allele and the third resolved both.

### Technical reproducibility

Technical replicates allowed us to assess the precision of ONT sequencing and Mahi-mahi genotyping. Forty-three sample–locus combinations from 20 VCGS samples were sequenced as two or three independent libraries. Restricting to the 39 sample–locus combinations in which every library was basecalled with the same model, replicate calls were identical for 56 of 70 alleles (80%; *ATXN1* in VCGS-53 was excluded because its libraries returned different numbers of alleles), and for alleles below 50 repeat units they were identical in 55 of 59 cases. The spread of repeat length calls increased with allele size, reaching a median of 7% of allele length above 300 repeats, based on four alleles (**Figure 4A, 4B)**. The four remaining combinations, each from a different sample, included libraries basecalled with the v4.3.0 and v5.2.0 models. In three of these, the expanded allele differed between libraries by 20% to more than two-fold, with the v4.3.0 call longer at *DMPK* in VCGS-04 and *ATXN8OS* in VCGS-65 and shorter at *C9orf72* in VCGS-27, which was called at 541 repeats from the v4.3.0 library and 1,208 repeats from the v5.2.0 library (**Figure 4C, Supplemental Figure 11)**. In each of these three, the v4.3.0 library was also sequenced with a different target file and had a shorter read N50, so the differences cannot be attributed to basecalling model alone. In the fourth, *FXN* in VCGS-02, both libraries used the same target file and the two models gave calls within 1% of each other (703 and 708 repeats).

**Figure 4.**
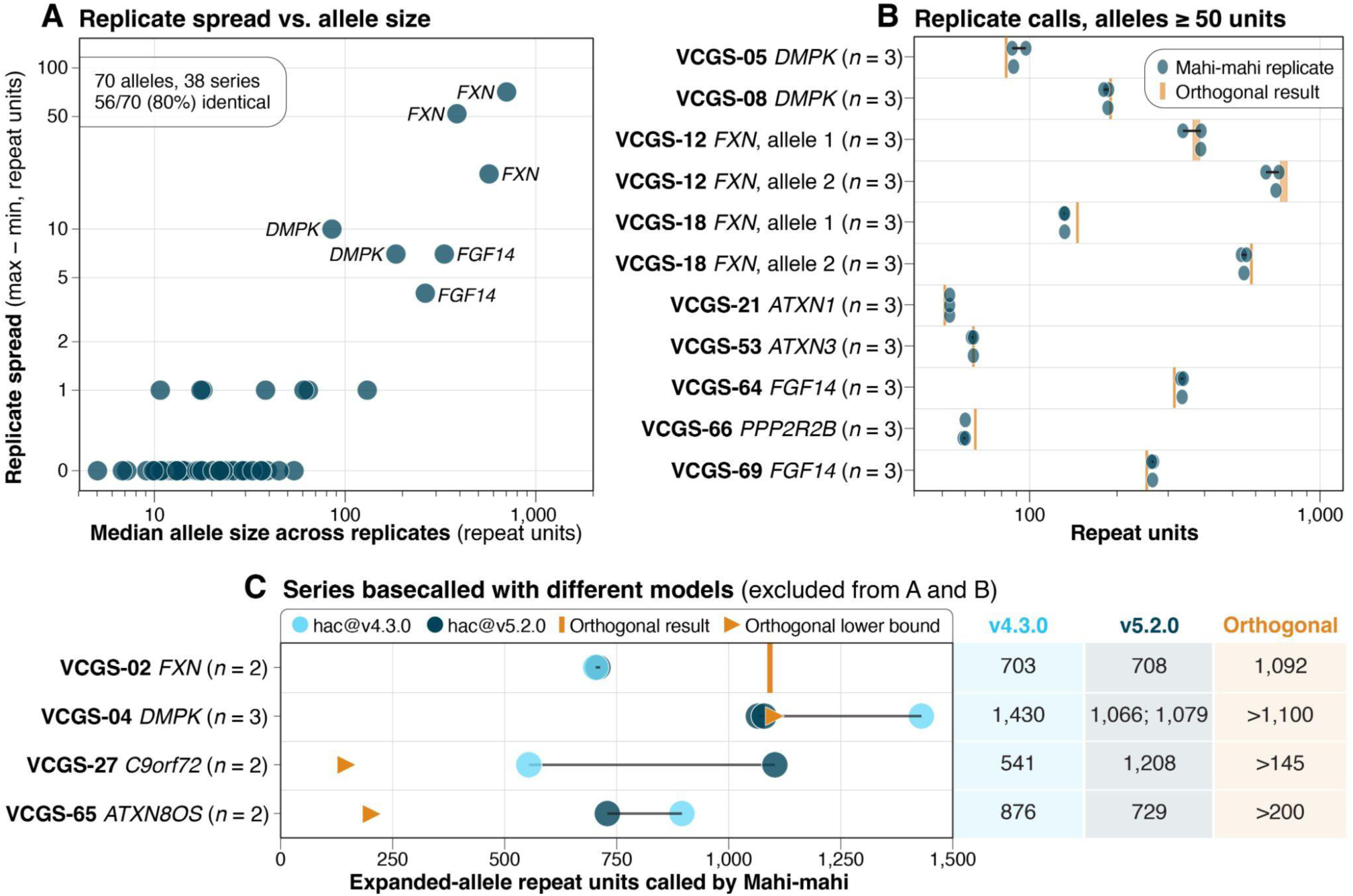
Technical reproducibility of Mahi-mahi genotypes. Replicate libraries were sequenced at VCGS from 20 samples, covering 43 sample–locus combinations. **(A)** Replicate spread (the largest minus the smallest call across libraries) plotted against the median allele size across replicates. The panel includes 70 alleles from the 38 replicate series in which every library was basecalled with the same model (HAC v5.2). *ATXN1* in VCGS-53 is excluded because its libraries returned different numbers of alleles. The *y*-axis is linear between 0 and 1 and logarithmic above 1. Alleles with a spread greater than one repeat unit are labeled by gene. **(B)** Individual replicate calls (circles) for every allele with a median size of 50 repeat units or more. Calls from the same allele are joined by a line, and *n* is the number of libraries. “Allele 1” and “allele 2” distinguish the two expanded alleles of samples with biallelic expansions. For VCGS-12, the orthogonal result was reported as ranges (350–400 and 730–770). **(C)** The four replicate series whose libraries were basecalled with different models (HAC v4.3 and HAC v5.2), excluded from A and B. Points show the expanded-allele call from each library, colored by model. The orthogonal result is marked as a tick when it was reported as an exact value and as a triangle when it was reported as a lower bound. The table at right lists each library’s call and the orthogonal result. Three of the four v4.3.0 libraries were also sequenced with a different target file and had a shorter read N50 than their v5.2.0 replicates, so these series cannot separate the effect of the model from those of target file and read length **(Supplemental Figure 11)**.

### Motif composition, interruptions, and basecalling artifacts

Mahi-mahi reports the motif composition and interruption structure of each allele directly from reads. Clinically, this is important at loci such as *RFC1*, where AAAGG expansions in the 1000 Genomes cohort, which are pathogenic only at very large sizes, overlapped pathogenic AAGGG expansions in size, so that motif as well as length determines their classification (Cortese et al. 2019) (**Figure 1D**). At *ATXN1*, where interruption of the CAG tract by CAT triplets modifies pathogenicity (Chung et al. 1993), Mahi-mahi reported CAT interruptions in UW-03 and none in the expanded allele of UW-04, matching the interruption status determined by SfaNI digestion of the PCR product in both samples (**Figure 5A,B**). At *BEAN1* in HG04094 from the 1000 Genomes cohort, Mahi-mahi showed that an allele of approximately 1,128 repeat units, reported by the assembly-based analysis as the reference motif, was composed mainly of TGAAA units rather than the reference TAAAA or the TGGAA insertion that causes SCA31 (**Figure 5E**). Finally, Mahi-mahi flags reads carrying systematic, motif-biased basecalling error. At *BEAN1* in the HG005 proband and mother, HG007, reads carrying strand-biased miscalls were flagged as suspicious, and the canonical genotype was returned (Sato et al. 2009); the mother carried two expanded alleles of which only one was transmitted to the proband, and the shared allele was sized consistently in both individuals **(Supplemental Figure 12)**. The same behavior was observed independently at *DAB1* in two 1000 Genomes samples, in which alleles called with a GGAAT rather than the canonical AAAAT motif were supported by reads flagged as suspicious (**Figure 5F, Supplemental Table 12)**.

**Figure 5.**
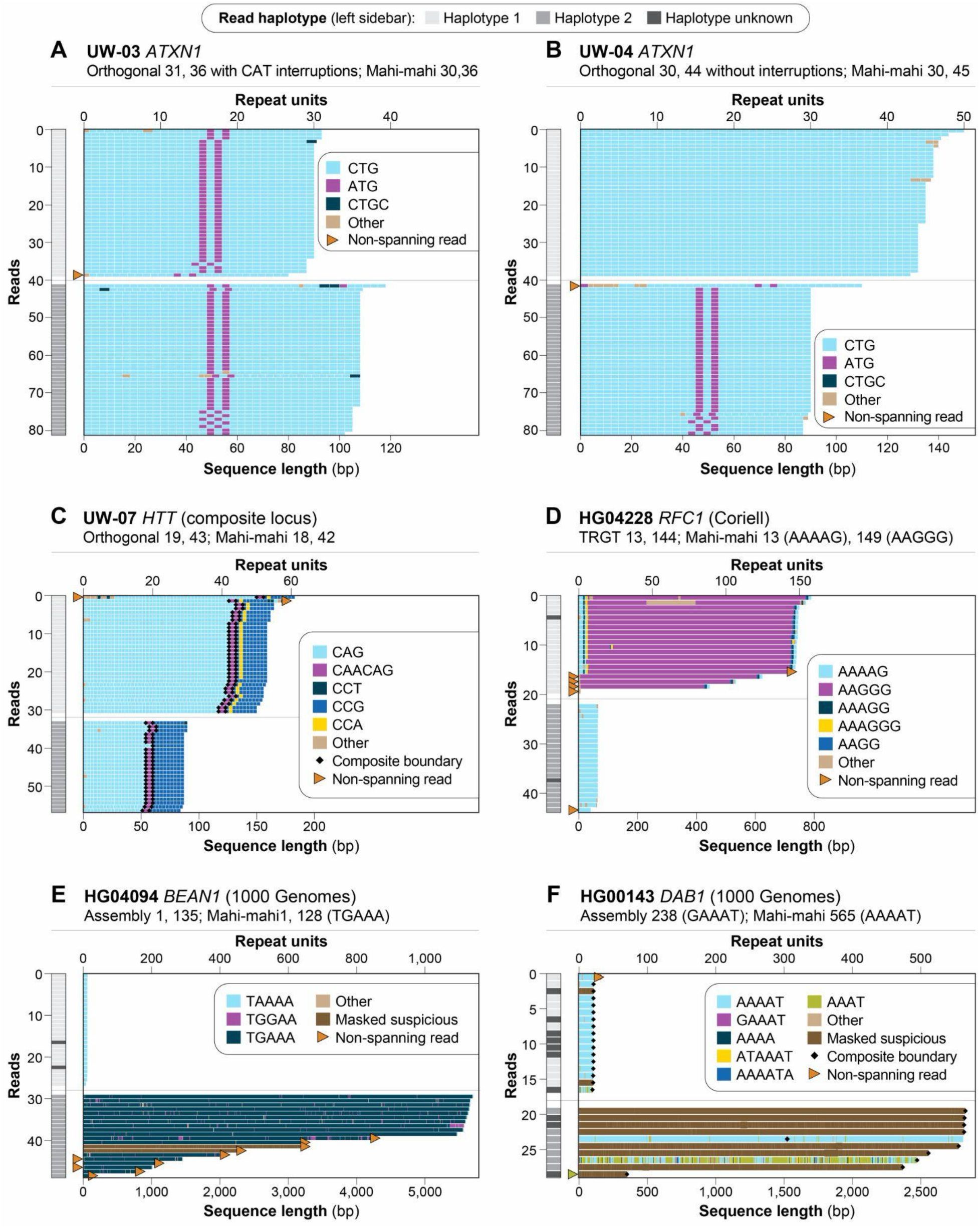
Read-level repeat structure resolved by Mahi-mahi. Each panel shows one sample and locus. Each row is a read, and reads are grouped by haplotype (sidebar). Colors indicate the motif assigned to each repeat unit, dark brown segments mark reads masked as suspicious, triangles mark reads that do not span the repeat, and black diamonds mark section boundaries. Panels A–C are UW samples sequenced by adaptive sampling and previously genotyped by orthogonal testing. **(A, B)** *ATXN1*, shown on the reverse strand so that CTG and ATG correspond to CAG and CAT. In UW-03 **(A)**, both alleles carry CAT interruptions, including the 36-repeat allele. In UW-04 **(B)**, the 45-repeat expanded allele is an uninterrupted CAG tract, and the normal allele carries two CAT interruptions. Both results match orthogonal testing. **(C)** *HTT* in UW-07. The CAG tract is followed by CAACAG and CCG units. A CCA unit between the CAACAG and CCG units is present on haplotype 1 only. **(D)** *RFC1* in Coriell line HG04228 (AS, v5.2 SUP). One allele is 13 units of the reference AAAAG motif. The other is about 149 units of the pathogenic AAGGG motif, preceded by a short AAAAG run. That is below the 400-unit threshold and consistent with carrier status (TRGT: 13, 144). **(E)** *BEAN1* in HG04094. The expanded allele (about 1,128 repeat units; 1,135 by assembly) consists mainly of TGAAA, not the reference TAAAA or the SCA31-associated TGGAA. Assembly-based analysis, which considers a fixed motif set, reported it as the reference motif. **(F)** *DAB1* in HG00143, a composite AAAAT/GGAAT locus. Assembly-derived calls reported a GAAAT tract in the expanded allele; on the opposite strand this would be the SCA37-associated ATTTC insertion. Mahi-mahi masked most expanded-allele reads, using them only for length estimation. Masked and unmasked reads together support an expansion of about 565 units of the canonical AAAAT motif.

## DISCUSSION

Here, we show that Mahi-mahi accurately genotypes tandem repeat expansions from nanopore sequencing data across reference samples, cell lines carrying known pathogenic expansions, and samples from three laboratories using different sequencing strategies. Using AS we targeted less than 2% of the genome and produced calls within one repeat unit of WGS for more than 99% of alleles in GIAB samples. At loci such as *HTT*, *ATXN1*, *DMPK*, and *PABPN1*, Mahi-mahi called the repeats within two repeat units of TRGT in every Coriell line; and in the cohorts from the three laboratories, Mahi-mahi detected an expanded allele in every sample with a pathogenic expansion reported by prior testing and assigned the same category (normal, intermediate, or pathogenic) as the referring laboratory for 98% of genotypes. Agreement on allele size below 100 repeat units was at the resolution of the PCR-based assays used for comparison, and independent libraries returned identical calls for nearly all alleles below 50 repeat units.

Most differences between Mahi-mahi and orthogonal methods did not reflect sizing error. Differences arose either because methods place the boundaries of a repeat differently, as at *FGF14*, *CNBP*, and *GOLGA8A*, or because they count repeat units differently: For composite loci, Mahi-mahi counts each contiguous section of a locus separately, including interruptions within it, whereas TRGT counts each motif wherever it occurs in the whole locus. Thus at *HTT* TRGT also counts the CAG within the adjacent CAACAG unit, and at *ATXN1* and *FMR1* Mahi-mahi includes the CAT or AGG interruptions that the TRGT motif count excludes. Offsets of this kind recurred against independent comparators: *HTT* calls were one repeat unit shorter than TRGT in both the GIAB and Coriell samples and than the orthogonal result in five of 23 alleles, and *FGF14* calls differed from both TRGT and orthogonal testing in every allele, in opposite directions, indicating that each method places the boundary of this repeat differently. *DMPK* was the exception, differing from orthogonal testing by five repeat units at four expanded alleles but not from TRGT, which points to clinical sizing at these alleles rather than to repeat definition. The remaining differences from TRGT arose because an allele contained a motif absent from the catalog entry for that locus, as at *RFC1*, *BEAN1*, *STARD7*, and *YEATS2*. Discrepancies of this kind are difficult to detect when tools are compared using summary statistics but are relevant at loci where a single repeat unit separates clinical categories, as illustrated by the *HTT* and *ATXN7* discordances described above. Similarly, the apparent discordance at *PABPN1* reflected the convention of reporting GCG triplets rather than the full GCN tract (Brais et al. 1998) **(Supplemental Figure 10)**. Harmonized locus definitions and reporting units, similar to what STRchive has begun to provide (Hiatt et al. 2025), will be necessary before results from different tools and laboratories can be compared directly, and we suggest that comparisons between tools report allele length in base pairs alongside motif counts so that definitional differences can be distinguished from sizing error.

Precision declined with allele size, as expected, and the largest alleles illustrate the limits of making a single size estimate when evaluating TR loci by directly sequencing them. For example, at *FXN* in Coriell lines, the repeat lengths of individual reads assigned to a single expanded allele spanned hundreds of repeat units, and TRGT reported similarly broad base-pair ranges at the same alleles; in laboratory samples, independent libraries of the largest *FXN* alleles differed by up to 13%. These observations are consistent with the somatic instability of large CTG and GAA expansions (Monckton et al. 1995; Long et al. 2017) and indicate that for such alleles the expansion is reproducibly detected while its size is better described as a range than as a single number. This is also how laboratories already report these alleles, typically as greater than a threshold, and every expansion of this size in our data was classified as pathogenic by every method. The basecalling model also affected the size of the largest alleles. When the same Coriell sequencing data were basecalled with the v5.2 and v6.0 HAC models, whole-genome calls for the expanded FXN allele differed by 79 to 325 repeat units in four of five lines, whereas calls at every other locus differed by no more than three repeat units between the two HAC models. The laboratory replicates basecalled with different models could not separate an effect of model from those of read length and target file, and the one such pair sequenced with the same target file agreed within 1%. We therefore recommend that implementations fix and report the basecalling model alongside the Mahi-mahi version and revalidate when either changes. Model choice also affects motif assignment, which matters more than length at loci where pathogenicity is defined by an inserted motif. In the 1000 Genomes data, assemblies built from reads basecalled with an earlier model reported GGAAT units at *DAB1*. Genotyped against the two common motifs in this locus (canonical: AAAAT, pathogenic: GAAAT), vamos reported these as GAAAT making them appear to be the SCA37 insertion, whereas Mahi-mahi masked the strand-biased reads carrying them and returned the canonical motif. Because the raw signal from archived clinical samples is often not retained and cannot be rebasecalled, read-level flagging of this kind is likely to be the main safeguard against reporting such artifacts, and we suggest that motif calls at insertion-defined loci such as *DAB1*, *BEAN1*, and the familial adult myoclonic epilepsy (FAME) loci (OMIM phenotypic series PS601068) be required to have support from reads on both strands.

Together, our results support the use of nanopore sequencing, and AS in particular, as a single test for repeat expansion disorders. Current clinical testing is typically ordered one locus at a time, with each assay contributing its own cost and turnaround time and a negative result at one locus prompting the next test (Depienne and Mandel 2021). Adaptive sampling requires no locus-specific reagents, which allows each laboratory to define its own target list and to add loci as new disorders are described without redesigning the assay (Stevanovski et al. 2022; Loose et al. 2016); indeed, the panels used here differed in the number of genes targeted yet shared a common set of repeat loci that was genotyped consistently across sites. Because the same data also capture single-nucleotide, structural, and phased variation across the targeted regions, a single AS run could potentially, after end user validation, address the differential diagnosis of a patient with ataxia or neuromuscular disease that would otherwise require several sequential tests (Rudaks et al. 2025). The same data also report allele-resolved methylation: at *FMR1*, Mahi-mahi recapitulated the expected pattern of X-chromosome inactivation in Coriell lines, and extending this to premutation and full-mutation alleles, where methylation status can inform interpretation, would address within the same assay a question that currently requires a separate methylation-sensitive test.

Our study has several limitations. First, comparison with clinical testing is limited by the precision of the orthogonal assays, which report sizes to within one or more repeat units at small alleles and only as a lower bound at large ones, so agreement at large alleles could be assessed only categorically. Second, although several tandem repeat genotypers support nanopore sequencing data (Mitsuhashi et al. 2019; Readman et al. 2021; De Coster et al. 2024; Ziaei Jam et al. 2024; Lougheed et al. 2026), we did not perform a systematic comparison with these tools here; such a comparison will require harmonized locus definitions for the reasons described above. Third, Mahi-mahi genotypes the loci defined in its catalog, and alleles composed of motifs absent from the catalog are sized but not counted in motif units, as observed at *BEAN1*, *STARD7*, and *YEATS2*. Fourth, alleles differing by a single repeat unit were not always resolved, and several loci were represented by few samples.

Overall, we find that Mahi-mahi accurately identifies and sizes repeat expansions from nanopore sequencing data, reports their motif composition and interruption structure, reports haplotype-specific methylation, and identifies reads affected by systematic basecalling error. Together with AS, it enables a single, scalable, and customizable test for repeat expansion disorders that performs comparably to the locus-specific clinical assays it could potentially replace in the future following full in-house validation by end users.

## CODE AVAILABILITY

Code used in this study can be found at https://github.com/millerlaboratory/2026_Mahi_mahi_preprint

## Data Availability

LRS data for 1KGP and GIAB samples are available online at https://s3.amazonaws.com/1000g-ont/index.html. Original sequencing data from laboratory samples cannot be shared due to institutional restrictions.

## ACKNOWLEDGEMENTS

DEM is supported by NIH grants DP5OD033357 and U24HG014453. Work conducted at VCGS was supported by the Victorian Medical Research Acceleration Fund (VMRAF). We thank Moustafa Shokrof and Tatiana Dvorkina for their work on early versions of Mahi-mahi, Edmund Day for generating datasets for testing and Miles Benton, Rocio Esteban, Bryan Leland, Priyesh Rughan, Rebecca Stubbs and all alpha testers for feedback on Mahi-mahi throughout development.

## ETHICS APPROVAL

Ethics approval for VCGS samples was obtained from the Royal Children’s Hospital Human Research Ethics Committee, Melbourne, Victoria, Australia (approval numbers RCH/HREC:125558 and RCH/HREC:28097). Samples analyzed at the University of Washington and UMC Utrecht were residual DNA specimens remaining after completion of diagnostic testing, which were used for test development and validation. These specimens were analyzed without identifying information, no additional material was collected from patients for this study, and in accordance with institutional policies governing the use of residual clinical specimens, review by an institutional review board or medical ethics committee was not required.

## USE OF AI-ASSISTED TOOLS

During preparation of this manuscript, the authors used Claude (Anthropic, Opus 5.5) to edit text for clarity and consistency, draft and revise figure legends, check numerical statements in the text against the supplemental tables, check references, and generate plotting code used to redraw and lay out figures. AI tools were not used to generate, modify or analyze sequencing data, to produce genotype calls, or to make analytic or interpretive decisions. All AI-assisted content was reviewed, verified and edited by the authors, who take full responsibility for the content of the publication.

## RESEARCH USE ONLY STATEMENT

Mahi-mahi and the Oxford Nanopore Technologies sequencing products evaluated in this study are for Research Use Only. Not for use in diagnostic procedures. This study reports research performance in the samples evaluated; it does not establish suitability for clinical diagnosis or patient management. References to disease-associated alleles, pathogenicity thresholds and prior clinical test results provide context for the research comparisons and do not constitute a recommendation for diagnostic use.

## AUTHOR CONTRIBUTIONS

Conceptualization: SJ, DEM, PR Software: SN, PR

Methodology: SBG, SL, DEM, SN, PR

Investigation: MA, JGB, RKC, JG, EJH, RJ, HMJ, SJ, RM, ALM, IvO, RNP, JR, SMR, DS, MT, AZ, MPGZ

Resources: JGB, KLIvG, PJL, SL, DEM, BvdZ

Formal analysis: MGE, SBG, SH, WK, TP, PR, SS

Supervision: KLIvG, SL, DEM, PR

Writing – original draft: SBG, HMJ, DEM, PR

Writing – review and editing: all authors

## COMPETING INTERESTS

RNP, RKC, SN, RM, AZ, SJ, HMJ, and PR are employees of Oxford Nanopore Technologies (ONT). UMC Utrecht has an ongoing research collaboration with ONT. KLIvG received travel support from ONT. SL has received travel support from ONT and Illumina. DEM is on the scientific advisory board at Inso Biosciences; is engaged in research agreements with ONT, PacBio, Illumina, and GeneDx; has received travel and/or research support from ONT, PacBio, and Illumina; holds stock options in MyOme and Inso Biosciences; and is a consultant for MyOme. The other authors do not have a competing interest.

