## Supplemental Figures for "Accurate repeat expansion genotyping from whole-genome and real-time targeted nanopore sequencing with Mahi-mahi"

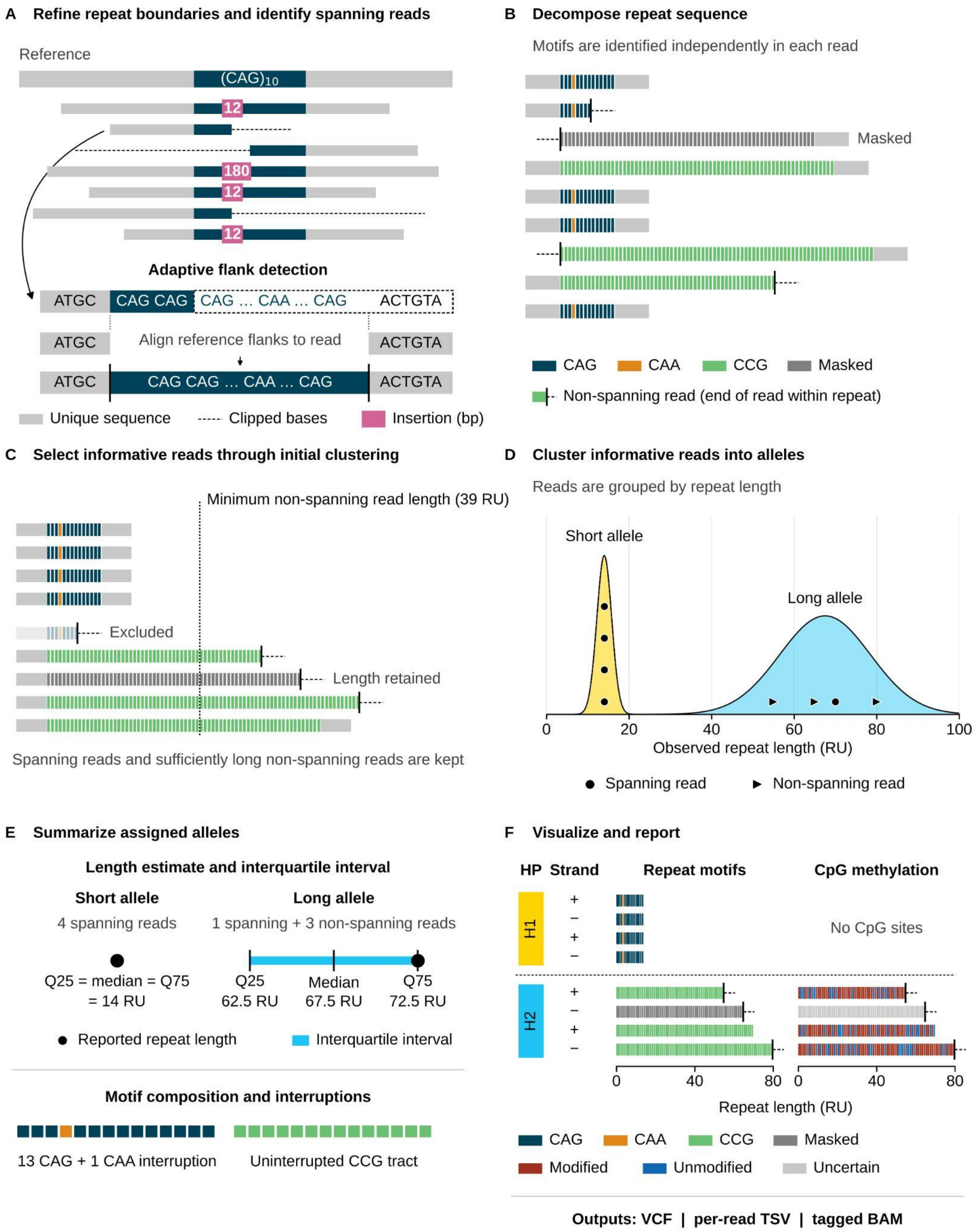

**Figure S1. Mahi-mahi workflow.**

**(A)** Reads overlapping each locus are retrieved, and the reference flanks are realigned to refine the repeat boundaries and to recover spanning reads from misaligned or soft-clipped alignments. **(B)** The repeat sequence of each read is decomposed into motifs independently, and reads whose motif composition is supported by only one strand are masked as suspicious. **(C)** Spanning reads, and non-spanning reads long enough to inform the length of the longest allele, are retained. **(D)** Informative reads are clustered into alleles by repeat length. **(E)** For each allele, the reported repeat length is the median of contributing reads, or the upper quartile when at least half are non-spanning, together with the interquartile range, motif composition, and interruptions. **(F)** Read-level visualizations show motif composition and CpG methylation for each read by haplotype and strand. Outputs are a VCF, a per-read TSV, and a tagged BAM file.

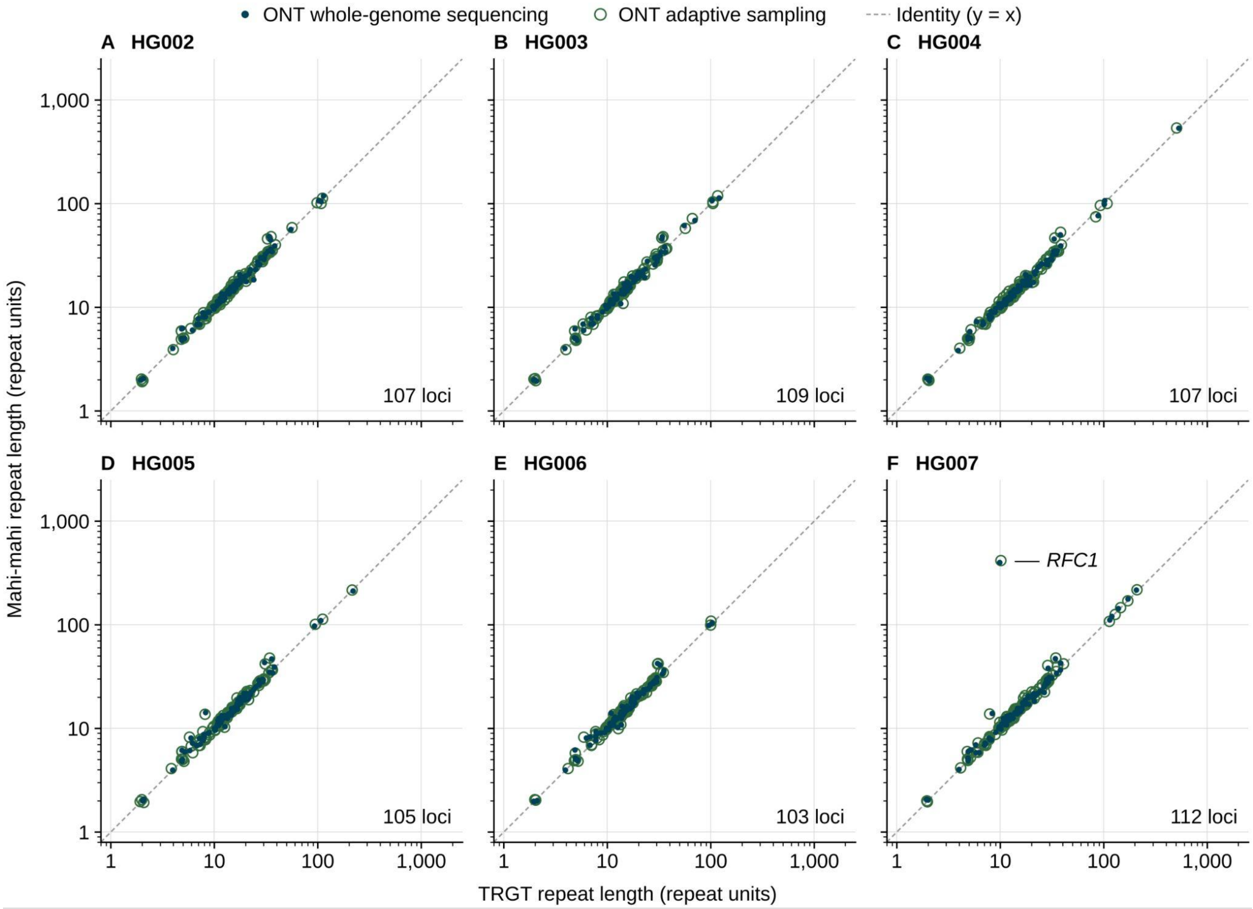

**Figure S2. Repeat length called by TRGT vs Mahi-mahi for HG002–7.**

ONT data were generated using either adaptive sampling (AS) or whole-genome sequencing (WGS). PacBio data for HG002-4 were collected on a Revio instrument while data for HG005-7 were collected on a Sequel II.

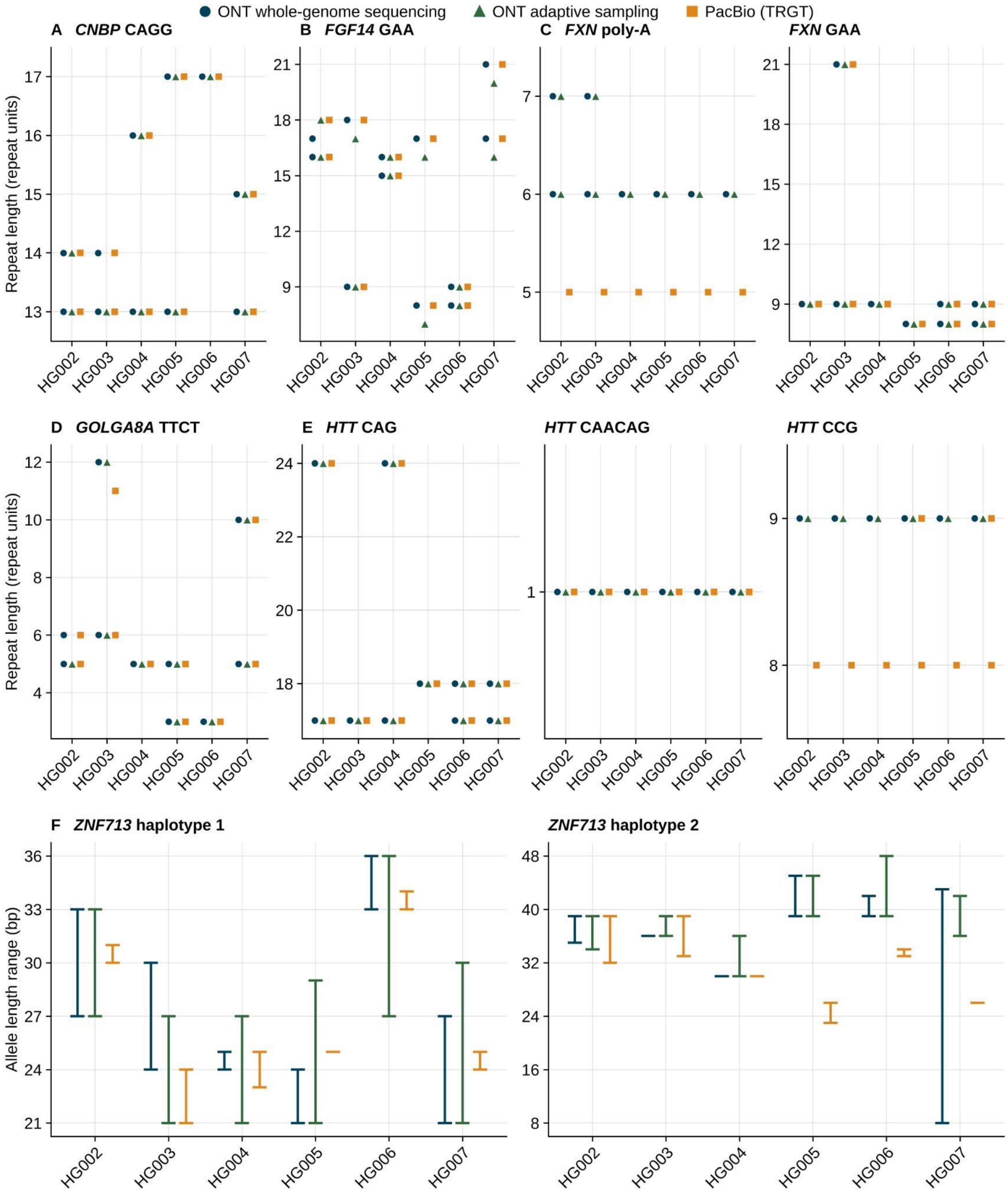

**Figure S3. Comparison of select loci from HG002-7.**

Mahi-mahi results are from WGS and AS, while PacBio data are from TRGT calls from Revio (HG002-4) or Sequel II data (HG005-7). **(A, B, D)** Mahi-mahi median count of the expanded motif (MRCN) versus TRGT motif count (MC) for the indicated motif at *CNBP*, *FGF14*, and *GOLGA8A*. HG004 and HG006 only have 1 allele genotyped by Mahi-mahi at *GOLGA8A*. **(C, E)**

Mahi-mahi repeat count for each component of the composite *FXN* and *HTT* loci, from the TSV output, versus TRGT MC for the same motif. If only one result or allele is shown then the individual was either homozygous or hemizygous for chrX variants. HG003 only has 1 allele genotyped by Mahi-mahi at *HTT*. **(F)** Allele length range in base pairs at *ZNF713* for each haplotype, from Mahi-mahi (ONT AS and WGS) and from TRGT (ALLR).

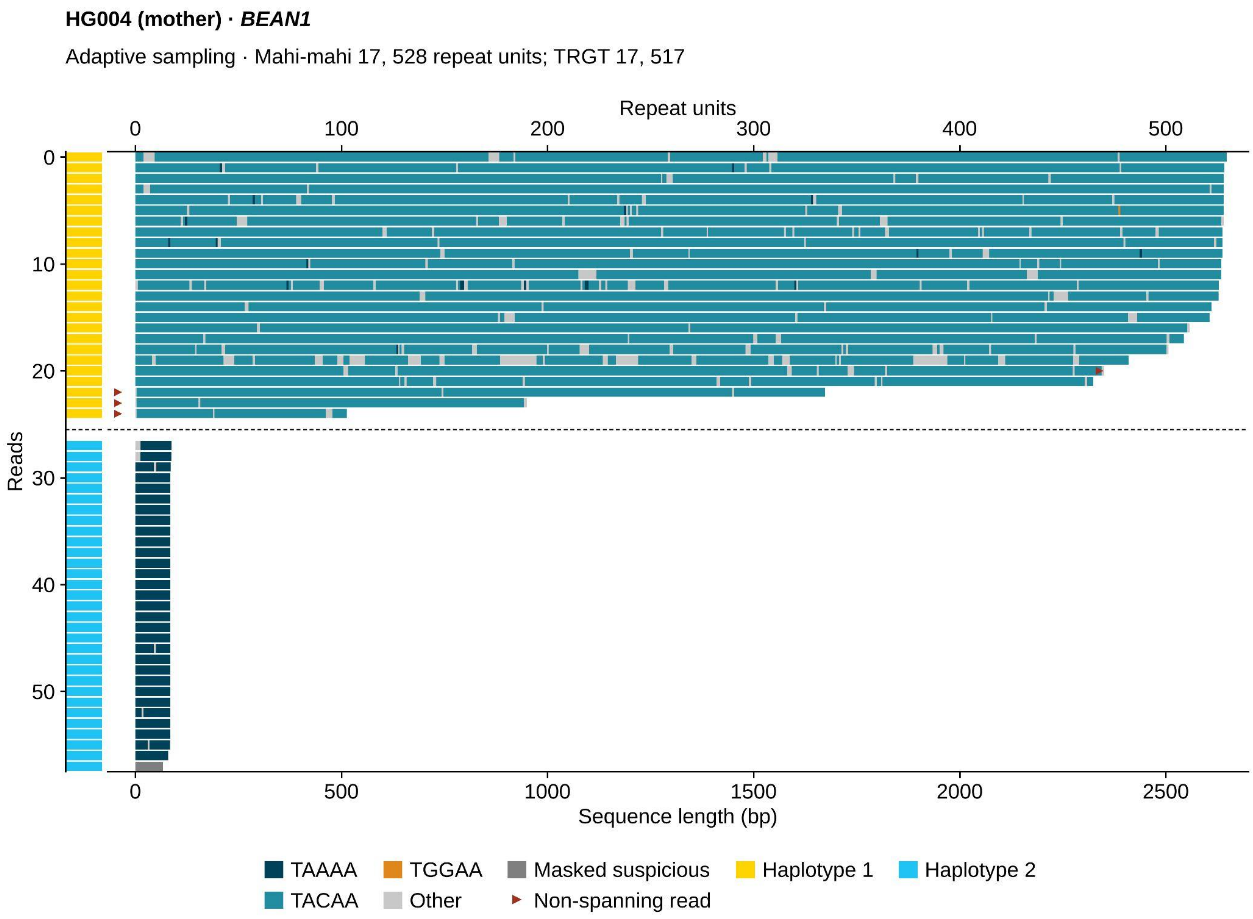

**Figure S4. A long nonpathogenic *BEAN1* allele in HG004.**

Mahi-mahi read-level visualization from adaptive sampling data, displayed as in Figure 5. HG004 carries a 17-unit TAAAA allele and a 528-unit allele composed almost entirely of TACAA, a motif absent from the catalog entry for this locus. TRGT sized the long allele at 517 units from its length in base pairs. A single TGGAA unit appears in one read; the pathogenic TGGAA expansion is not present.

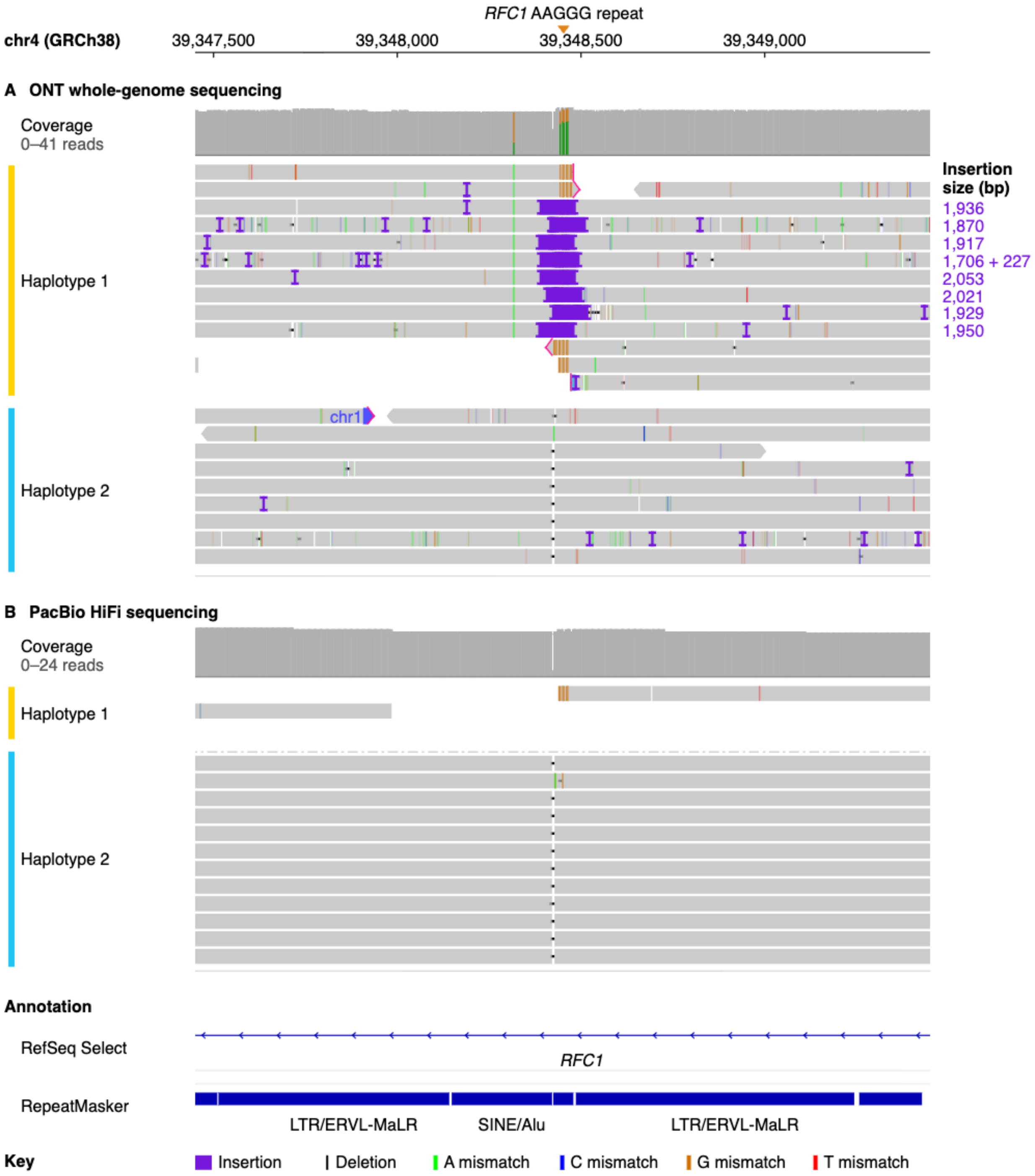

**Figure S5. An expanded *RFC1* allele is not present in the PacBio Sequel II data from HG007.**

IGV screenshot is from chr4:39,345,739-39,351,232 (hg38) and shows data at *RFC1* in HG007; reads are grouped by phase. In the ONT data (top), reads on haplotype 1 carry insertions of up to approximately 2 kb at the repeat, whereas in the PacBio data (bottom) the only haplotype 1 read that reaches the repeat ends there and does not extend into the expansion. This is consistent with random undersampling of the expanded allele in the PacBio data.

**Figure S6. The three largest discordances between Mahi-mahi and assembly-derived vamos calls in 1000 Genomes samples.**

Each group of panels shows one sample–locus combination on its own page: *RFC1* in HG04211 **(A–C)**, *RFC1* in HG00253 **(D–F)**, and *YEATS2* in HG04033 **(G–I)**. In each case, Mahi-mahi called one allele that agreed with vamos and one expanded allele that vamos did not report. **(A, D, G)** Mahi-mahi clustering of reads into alleles. Each point is a read positioned by its repeat length in bases, colored by the cluster to which it was assigned and shaped by haplotype; the blue curve is the density estimate, dashed lines mark detected peaks, and solid black lines mark the final allele clusters. **(B, E, H)** Alignments to GRCh38 of both assembly haplotypes and of raw reads basecalled with the v6.0 HAC model and grouped by haplotype, with the Mahi-mahi and vamos repeat intervals marked below. Insertion labels give the alignment difference relative to GRCh38, not the total repeat length. **(C, F, I)** Mahi-mahi read-level visualizations of motif composition, in which each row is a read, colors indicate motifs, gray segments mark reads masked as suspicious, red markers indicate reads that do not span the repeat, and the sidebar indicates haplotype.

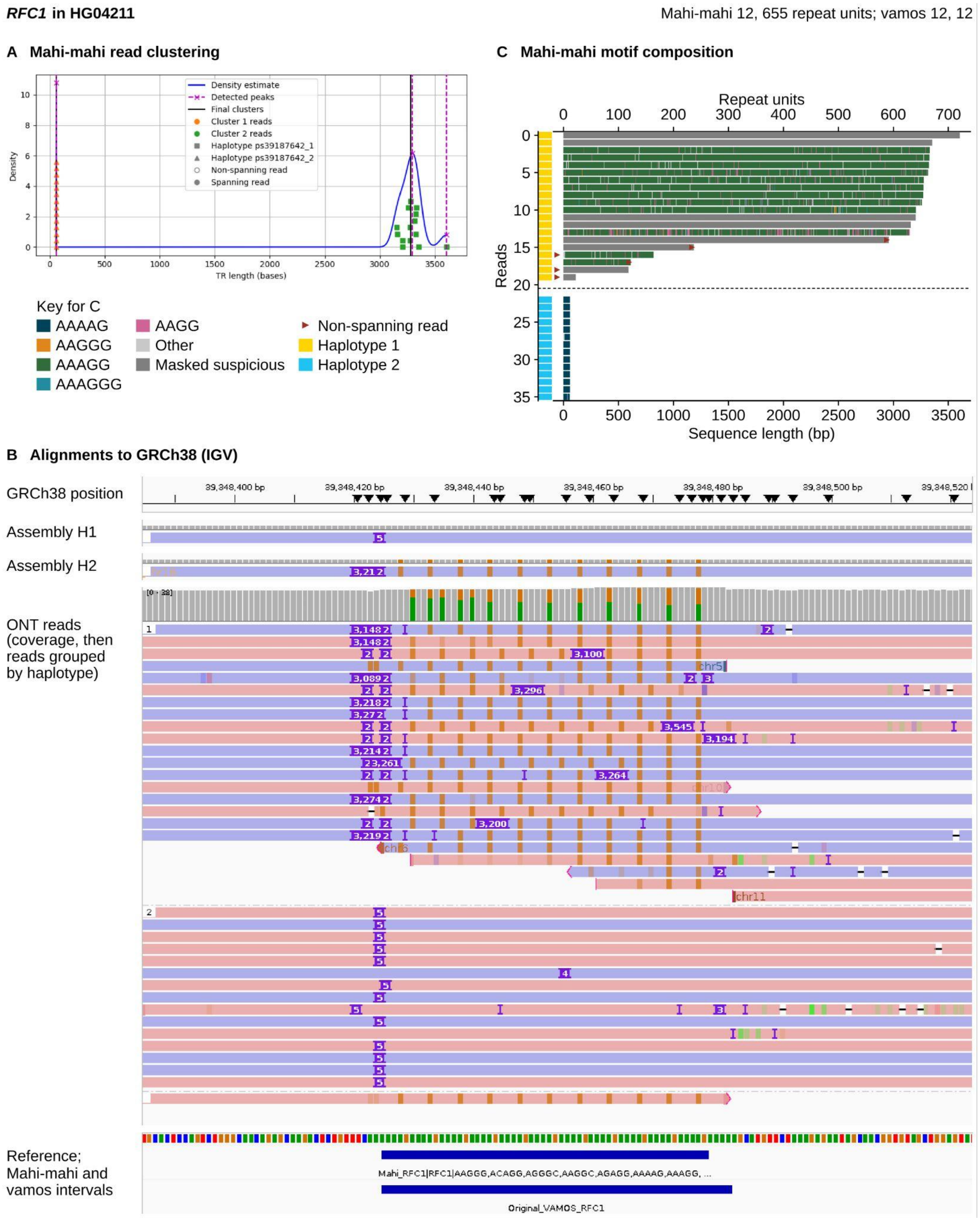

**S6, A–C.** At *RFC1* in HG04211, Mahi-mahi called alleles of 12 and 655 repeat units, whereas vamos called 12 and 12. One assembly haplotype carries a 3,212-bp insertion that is supported by the raw reads but was placed just upstream of the vamos interval during alignment, so it was not included in the vamos call.

***RFC1* in HG00253**

**D Mahi-mahi read clustering**

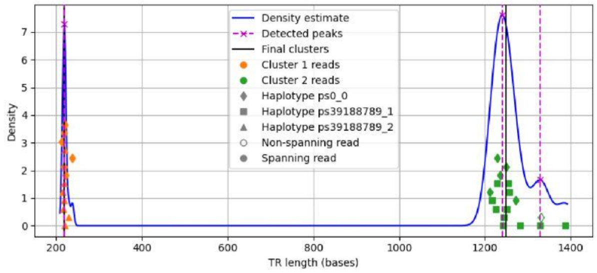

**F Mahi-mahi motif composition**

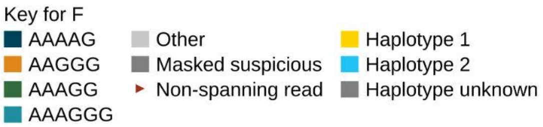

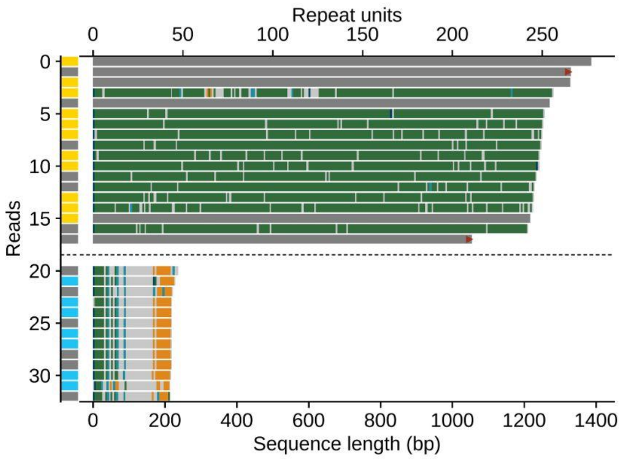

**E Alignments to GRCh38 (IGV)**

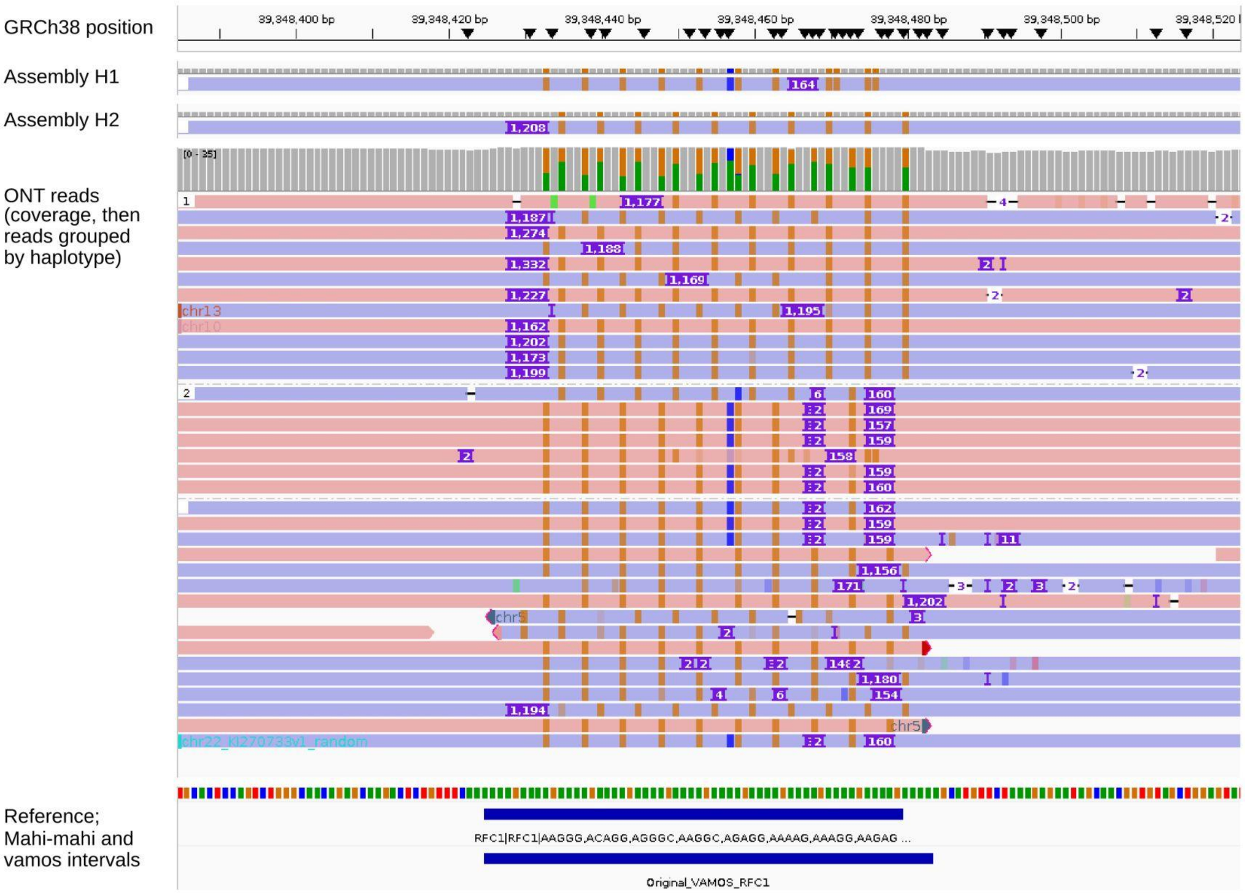

**S6, D–F.** At *RFC1* in HG00253, Mahi-mahi called alleles of 44 and 250 repeat units, whereas vamos called 45 for both. The assembly haplotypes carry insertions of 164 and 1,208 bp, and the raw reads separate into corresponding short and long groups. Here the larger insertion lies within the vamos interval, and the reason vamos did not report the expanded allele could not be determined.

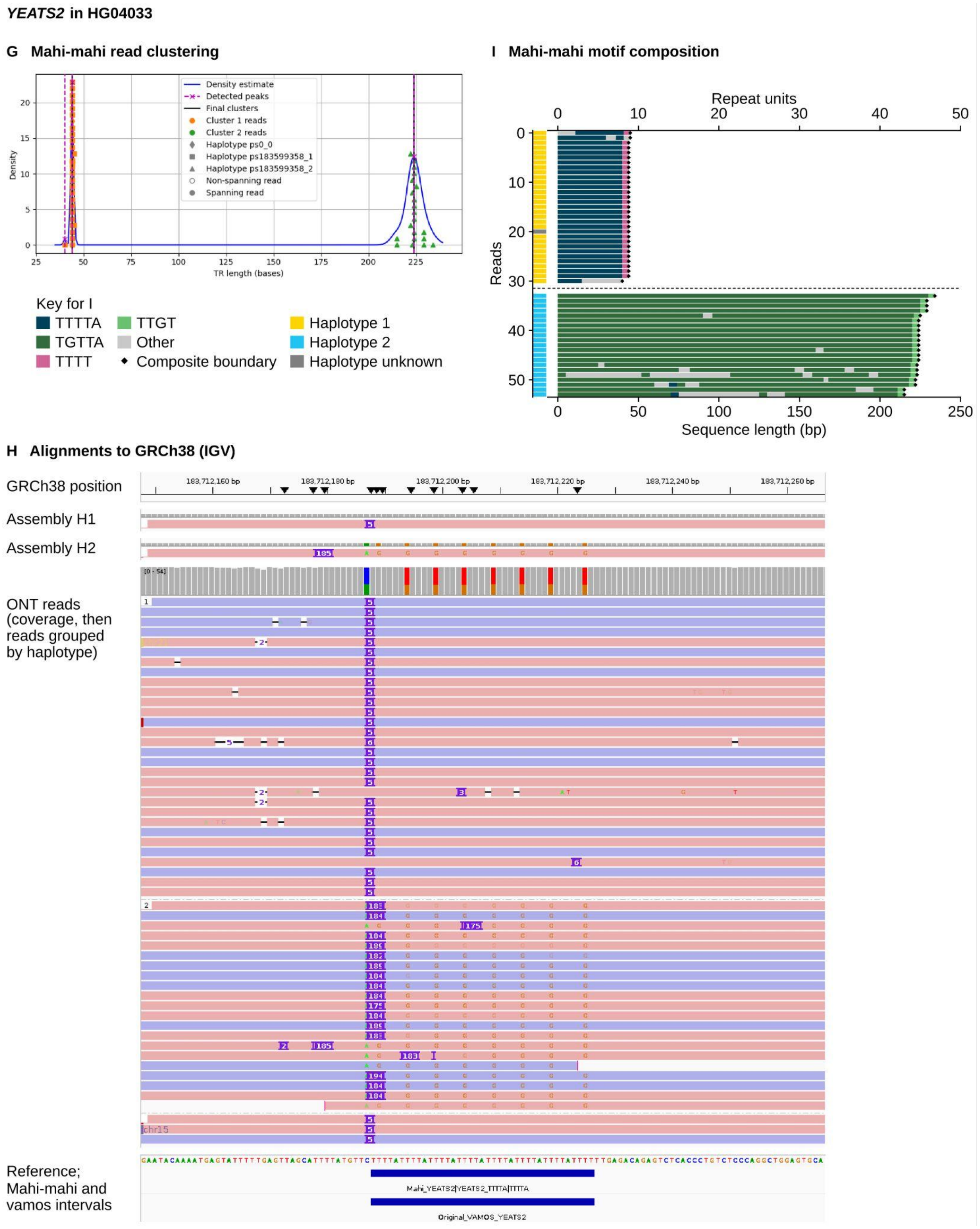

**S6, G–I.** At *YEATS2* in HG04033, Mahi-mahi called alleles of 9 and 45 repeat units, whereas vamos called 9 and 8. One assembly haplotype carries a 185-bp insertion composed largely of TGTTA units that was placed upstream of the vamos interval, so vamos called that allele at close to reference length. The allele was not removed by the filter for reference-matching sequence because it carries T>G substitutions relative to GRCh38. The raw reads and the Mahi-mahi visualization of this composite locus both support the longer allele.

**Figure S7. Expanded alleles identified by Mahi-mahi in 1000 Genomes samples that were not reported in the assembly-based analysis.**

Each sample is shown on its own page in three panels. The first panel shows Mahi-mahi clustering of reads into alleles. Each point is a read positioned by its repeat length in bases, colored by the cluster to which it was assigned and shaped by haplotype; the blue curve is the density estimate, dashed lines mark detected peaks, and solid black lines mark the final allele clusters. The second panel shows alignments to GRCh38 of both assembly haplotypes and of raw reads basecalled with the v6.0 HAC model and grouped by haplotype, with the Mahi-mahi and vamos repeat intervals and the lengths reported by each tool marked below. Insertion labels give the alignment difference relative to GRCh38, not the total repeat length. The third panel shows the Mahi-mahi read-level visualization of motif composition, displayed as in Figure 5.

***TCF4* in HG00231** Mahi-mahi 11, 267 CTG repeats

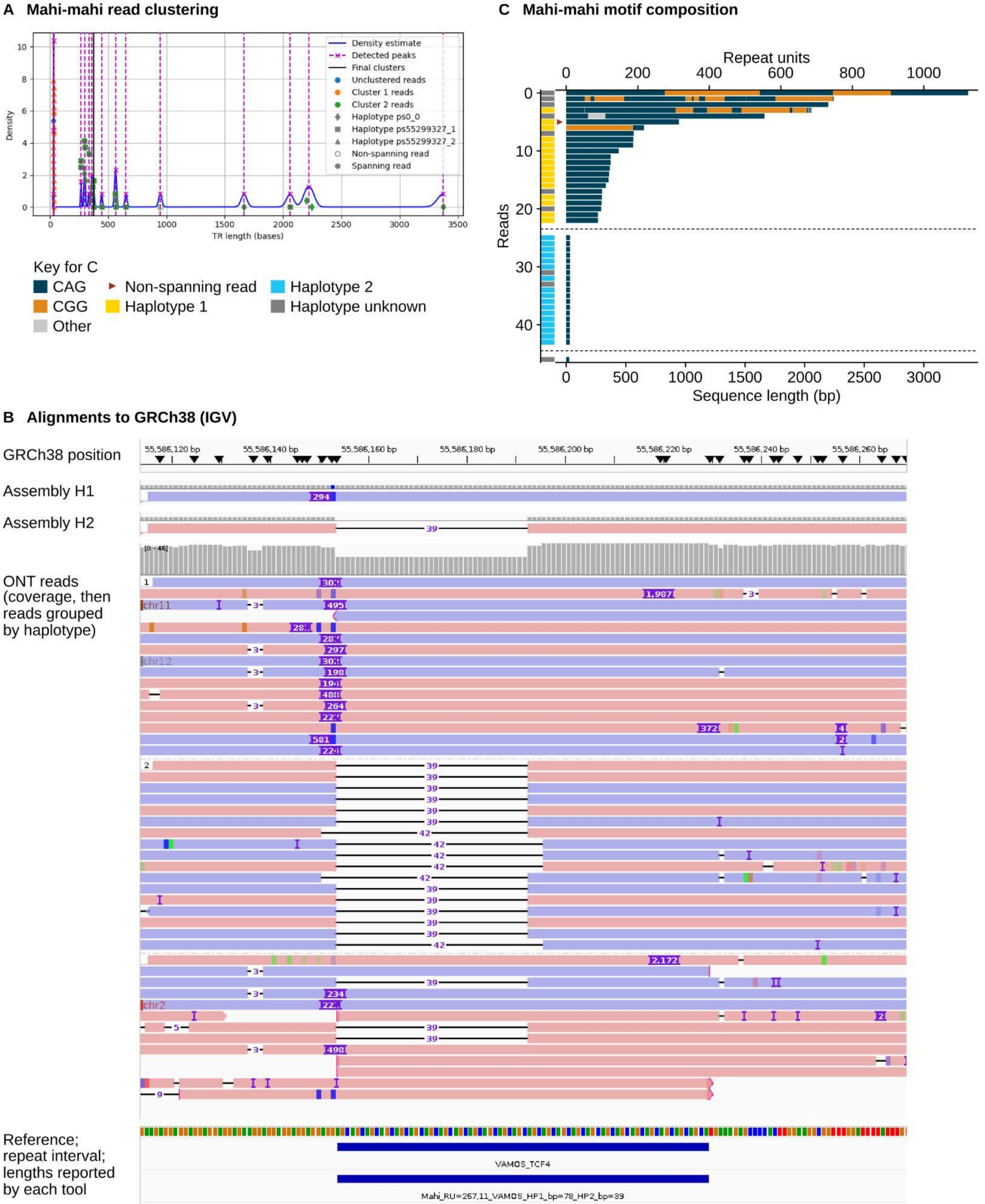

**S7, A–C:** *TCF4* in HG00231. Mahi-mahi called alleles of 11 and 267 CTG repeats. One assembly haplotype carries a 294-bp insertion placed just upstream of the vamos interval, so the vamos call for that haplotype was close to reference length. The allele was therefore removed from the concordance analysis by the filters designed to exclude assembly errors. Reads from the expanded allele are markedly heterogeneous in length, ranging from approximately 250 to more than 3,000 bases **(A, C)**.

***TCF4* in HG02351** Mahi-mahi 16, 100 CTG repeats

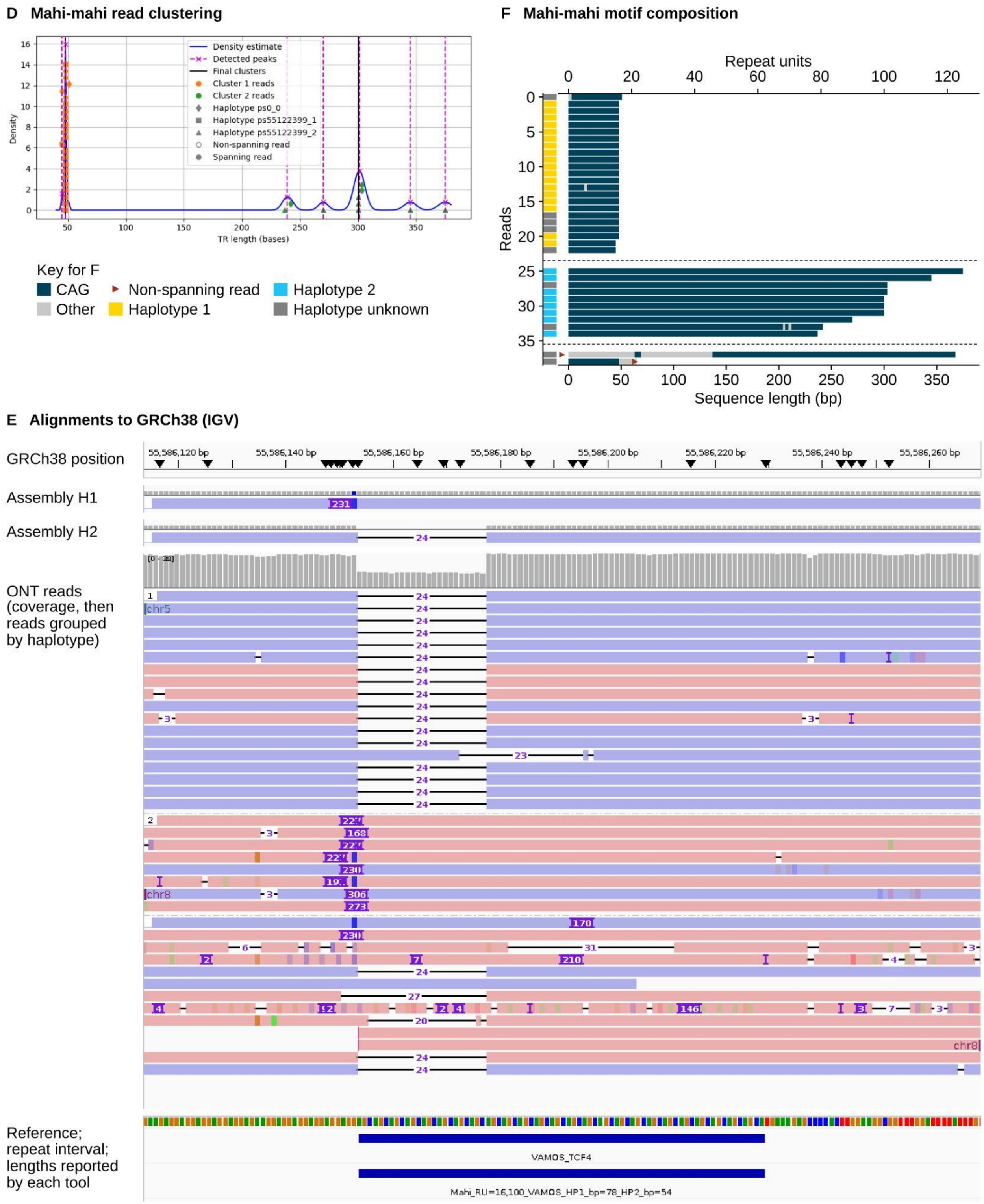

**S7, D–F:** *TCF4* in HG02351. Mahi-mahi called alleles of 16 and 100 CTG repeats. As in HG00231, a 231-bp insertion in one assembly haplotype was placed upstream of the vamos interval, and the allele was removed by the same filters. The expanded allele is more uniform in length than in HG00231 **(D)**.

***FXN* in HG03757** Mahi-mahi 8, 65 repeat units

**G Mahi-mahi read clustering**

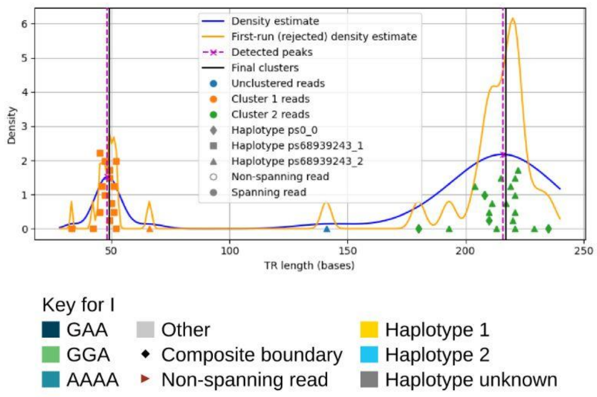

**I Mahi-mahi motif composition**

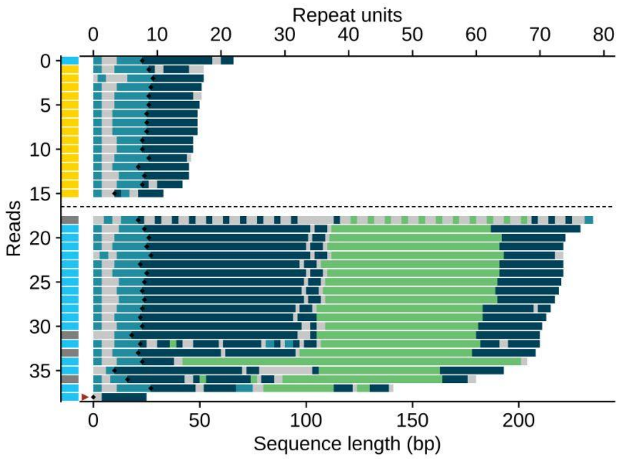

**H Alignments to GRCh38 (IGV)**

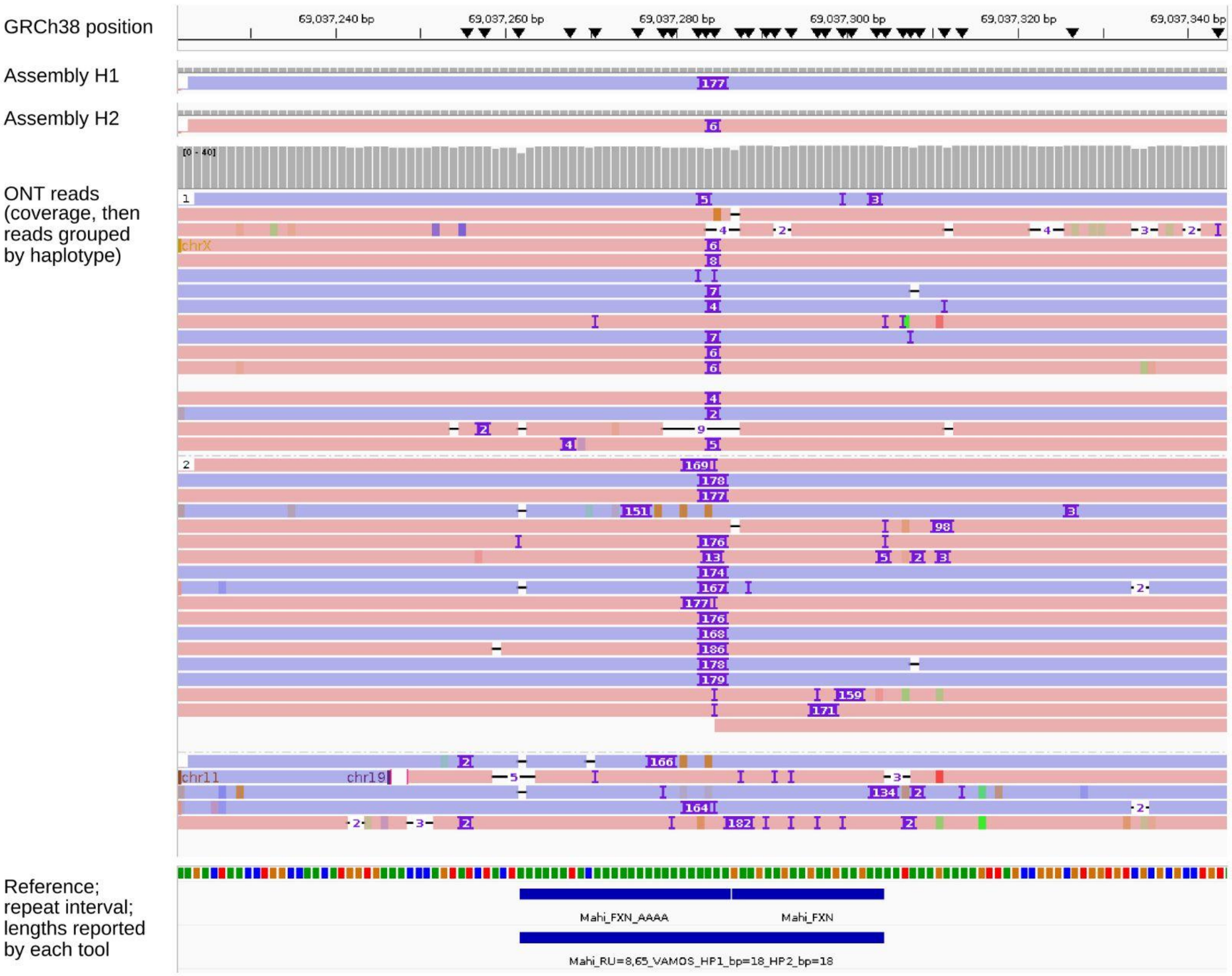

**S7, G–I:** *FXN* in HG03757, which Mahi-mahi treats as a composite of the adjacent poly-A tract and the GAA repeat, with black diamonds marking the boundary between them **(I)**. Mahi-mahi called alleles of 8 and 65 repeat units, whereas vamos reported reference length for both haplotypes despite a 177-bp insertion in one assembly haplotype **(H)**. The expanded allele is a composite in which tracts of GAA triplets (dark blue) are separated by a similar number of GGA triplets (light green) **(I)**. An initial density estimate (orange) was rejected and the reads were re-clustered **(G)**.

***FXN* in HG04235** Mahi-mahi 8, 63 repeat units

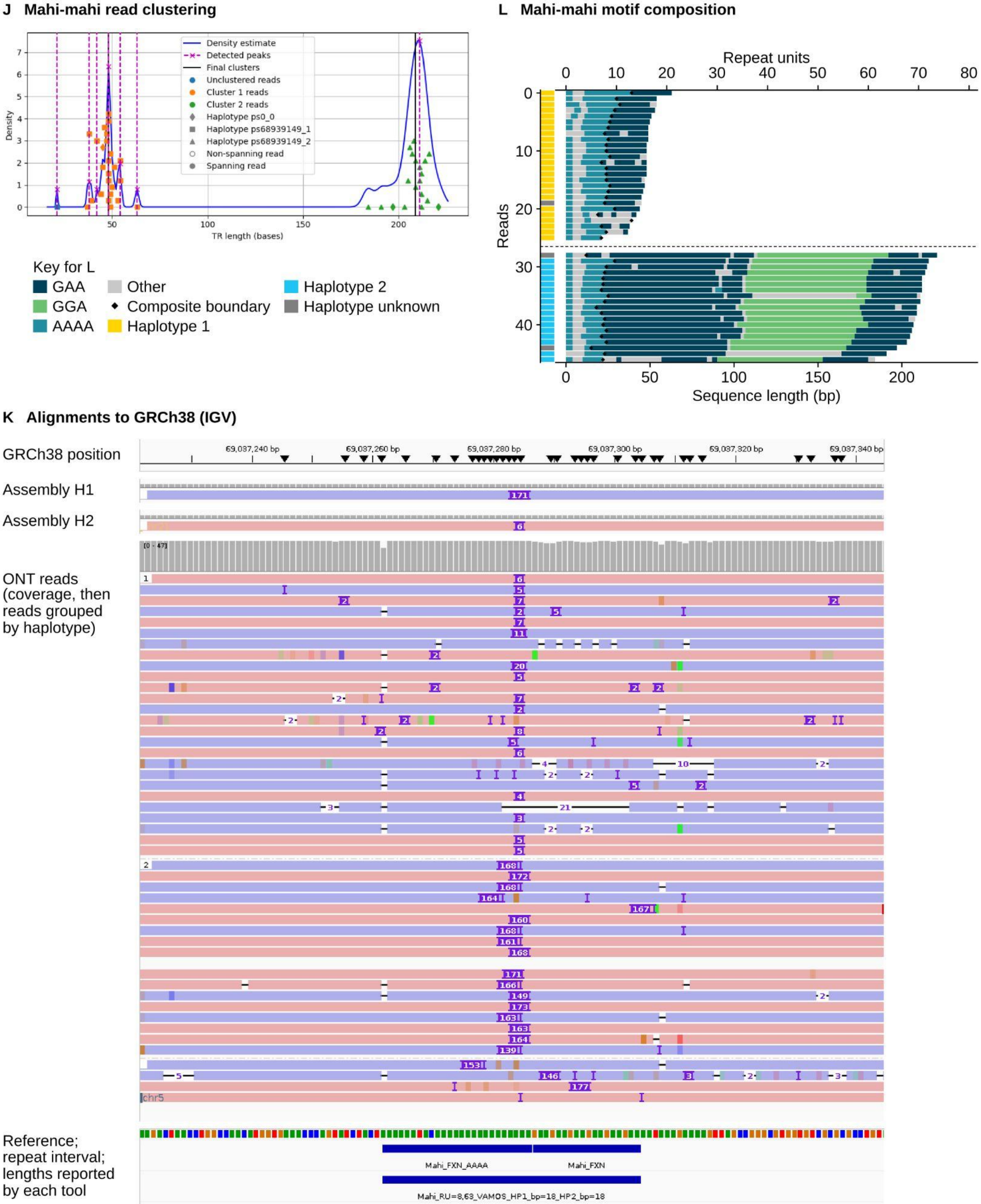

**S7, J–L:** *FXN* in HG04235. Mahi-mahi called alleles of 8 and 63 repeat units. vamos again reported reference length despite a 171-bp insertion in one assembly haplotype **(K)**, and the expanded allele has the same GAA–GGA composite structure **(L)**.

***FGF14* in HG00556** Mahi-mahi 10, 611 repeat units; vamos long allele 1,832 bp

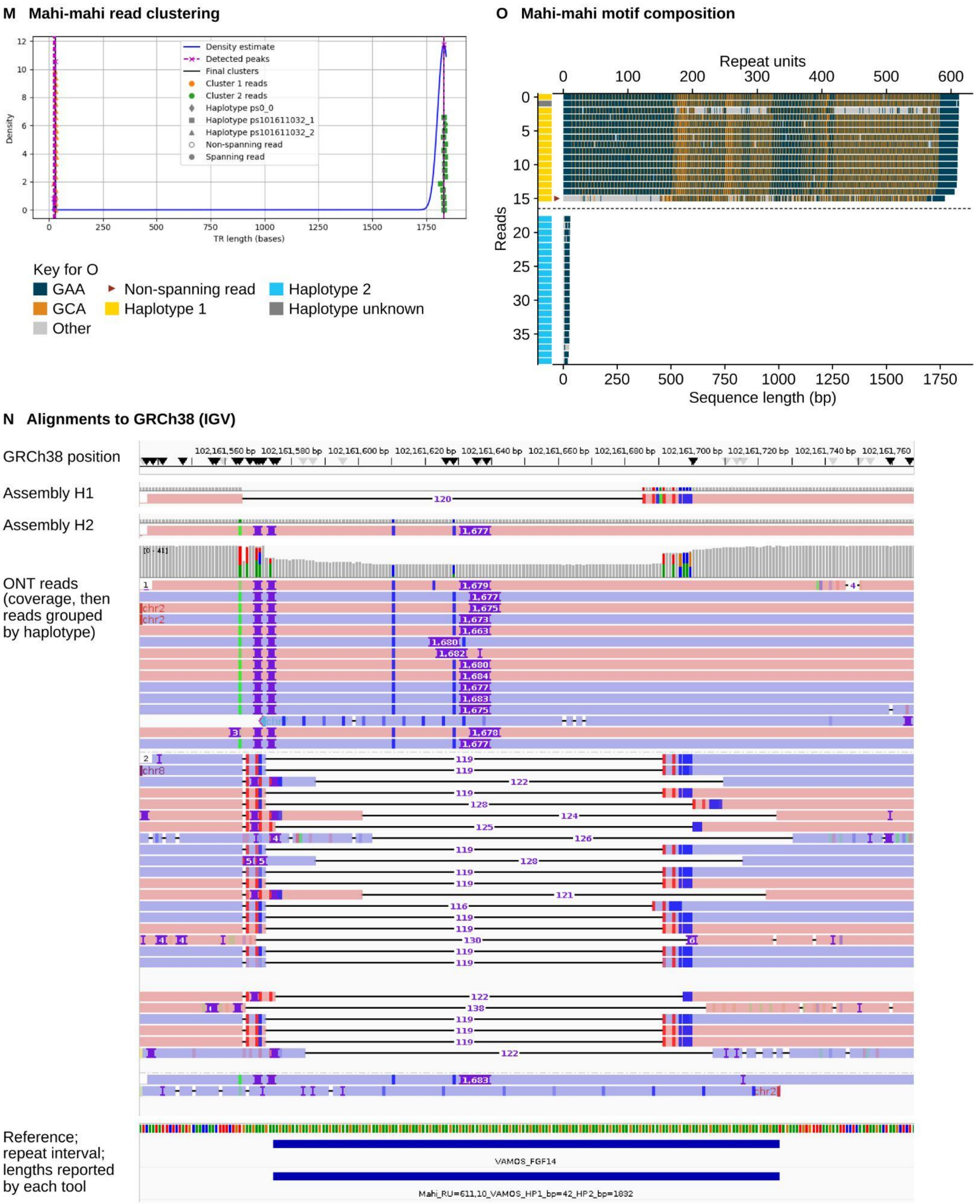

**S7, M–O:** *FGF14* in HG00556. Mahi-mahi called alleles of 10 and 611 repeat units, consistent with the vamos length of the long allele (1,832 bp) **(N)**. The expanded allele is interrupted throughout by GCA units **(O)**.

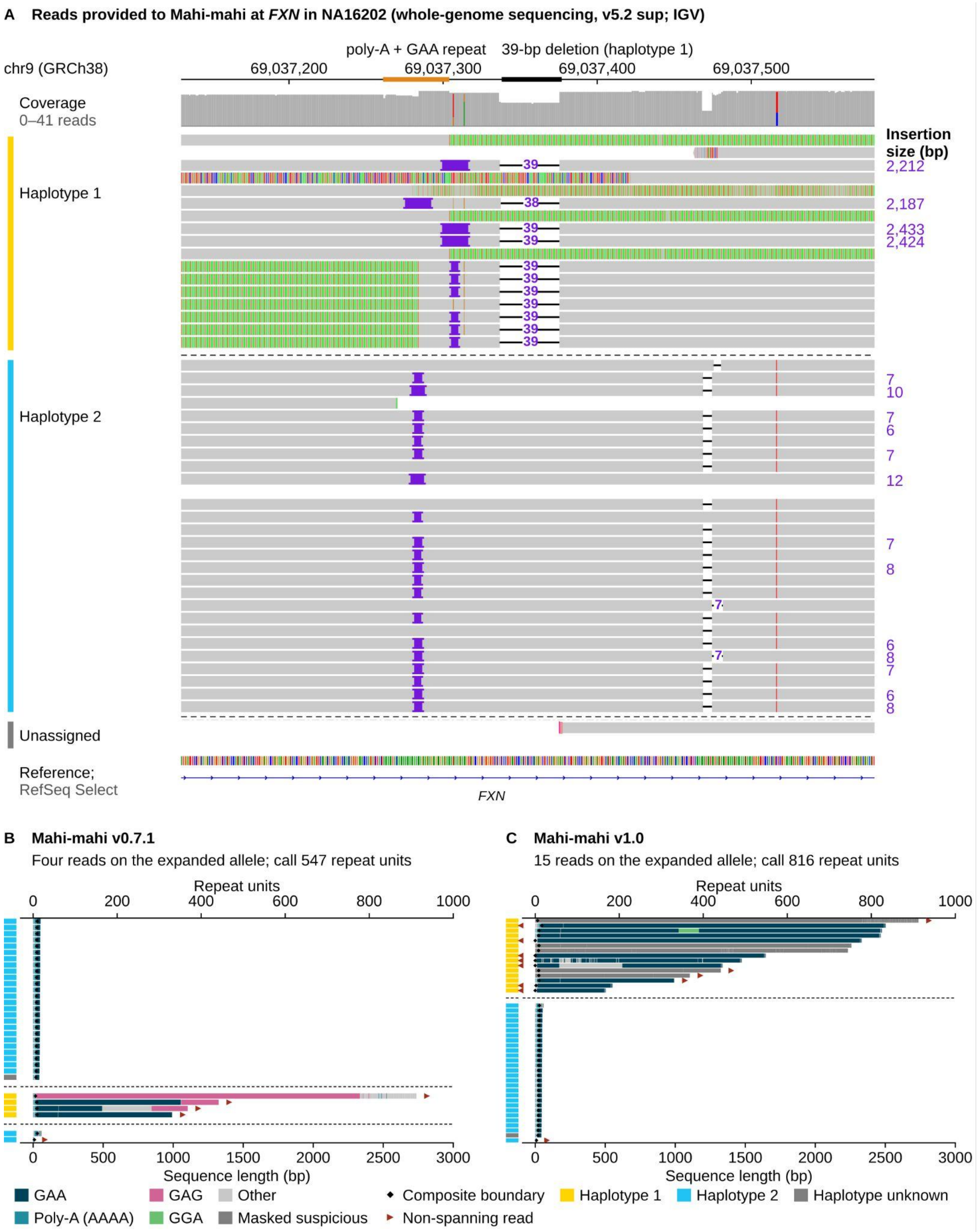

**Figure S8. Handling of a flanking deletion at the expanded *FXN* allele in NA16202**

Whole-genome sequencing data basecalled with the v5.2 SUP model. **(A)** IGV view of the haplotagged alignments from EPI2ME wf-human-variation, which are the reads provided to Mahi-mahi, grouped by haplotype. On the expanded haplotype (haplotype 1), reads carry insertions of approximately 2.2–2.4 kb at the repeat and a 39-bp deletion in the downstream flank; many reads are soft-clipped at the repeat (colored bases) rather than aligned through it. Insertion sizes are shown at right and give the alignment difference relative to GRCh38. **(B)** Mahi-mahi v0.7.1 assigned only four reads to the expanded allele, all non-spanning, giving a call of 547 repeat units. **(C)** Mahi-mahi v1.0, which accommodates deletions in the flanking sequence, assigned 15 reads to the expanded allele and called it at 816 repeat units, close to the Coriell-reported size of approximately 830. Read-level visualizations are displayed as in Figure 5.

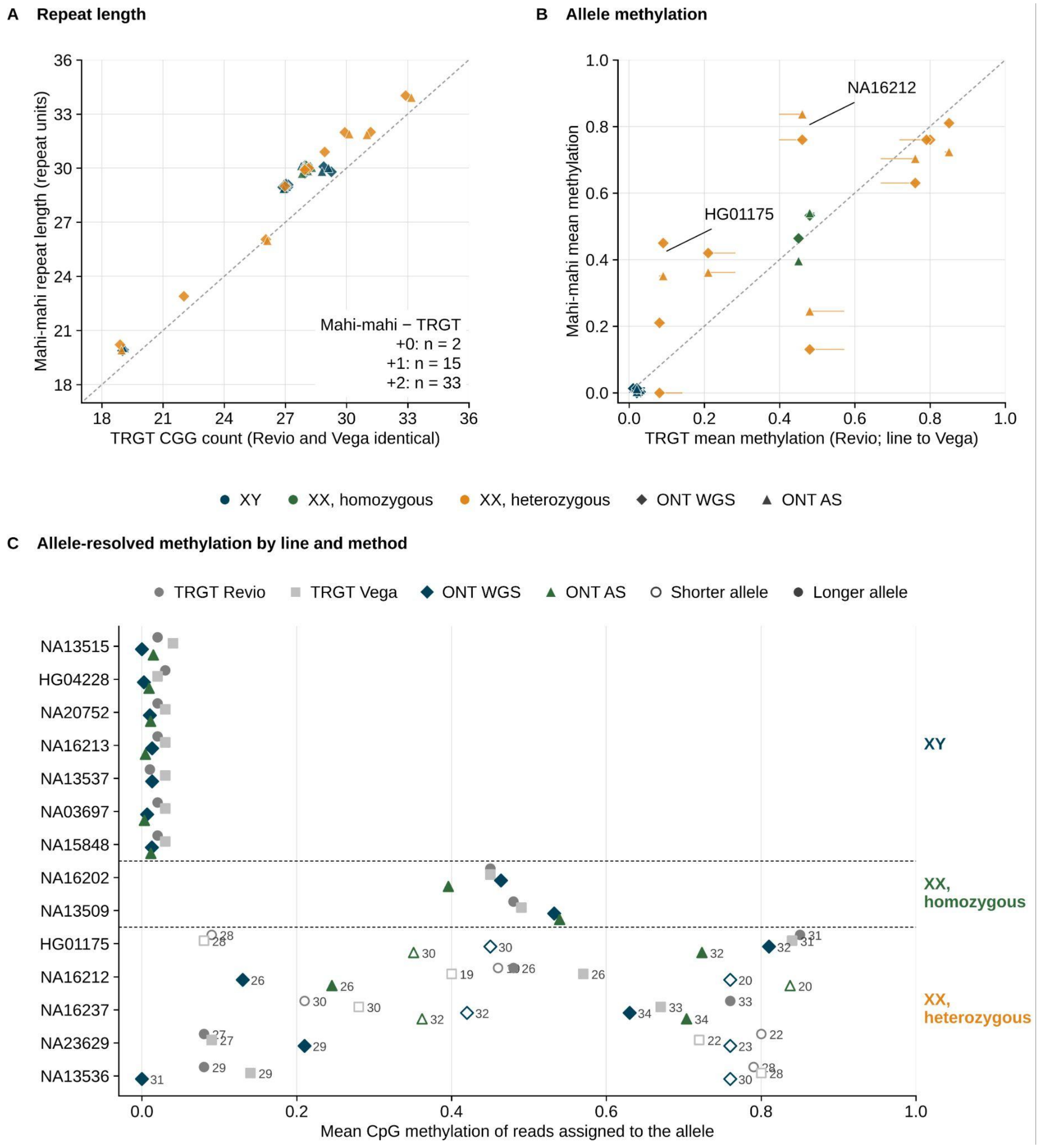

**Figure S9.** ***FMR1*** **repeat length and methylation in Coriell lines.**

**(A)** Mahi-mahi repeat length from ONT whole-genome sequencing (diamonds) and adaptive sampling (triangles) versus TRGT CGG count from PacBio PureTarget data, which were identical between Revio and Vega instruments. Points are colored by sex-chromosome complement (XY or XX) and zygosity and jittered slightly to reduce overlap. **(B)** Mahi-mahi versus TRGT mean CpG methylation for each allele; horizontal lines connect the Revio and Vega estimates. **(C)** Mean methylation of each allele by line and method. For heterozygous XX lines, open symbols mark the shorter allele and filled symbols the longer, with allele length labeled; for XY lines and homozygous XX lines, a single value is shown per method. TRGT reports two alleles at *FMR1* in XY lines; these are identical and are shown once. Adaptive sampling data were not available at *FMR1* for NA13537, NA23629, and NA13536.

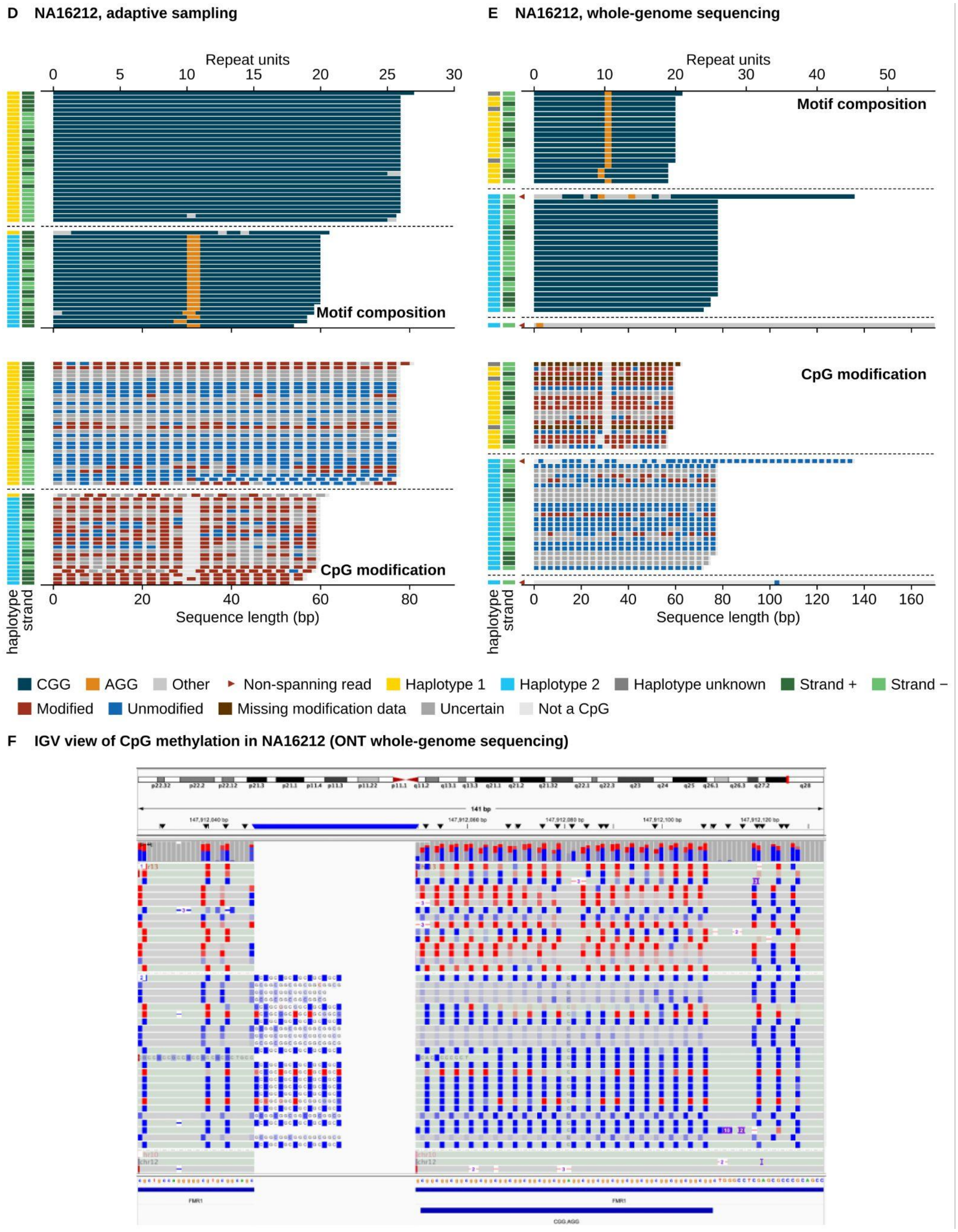

**S9, (D)** Mahi-mahi read-level visualizations of *FMR1* in NA16212 from adaptive sampling data, the sample in which Mahi-mahi and TRGT methylation estimates differed. The upper plot shows motif composition (CGG, navy; AGG, orange) and the lower plot shows the modification call at each CpG in the same reads (modified, red; unmodified, blue; missing modification data, brown; uncertain, dark gray; not a CpG, light gray). Each row is a read, and sidebars indicate haplotype (yellow, haplotype 1; cyan, haplotype 2) and strand (dark green, +; light green, −). Reads from the 20-repeat allele, which carries a single AGG interruption, are predominantly methylated (mean 0.84), and reads from the uninterrupted 26-repeat allele are predominantly unmethylated (0.25), whereas TRGT reported similar methylation for both alleles (0.46 and 0.48 on Revio). Alleles are matched by length. **(E)** As in D, from whole-genome sequencing data. Reads from the 20-repeat allele with its AGG interruption are again predominantly methylated (0.76) and reads from the 26-repeat allele predominantly unmethylated (0.13), reproducing the allele-specific pattern seen with adaptive sampling. **(F)** IGV view of CpG methylation in NA16212 at *FMR1*.

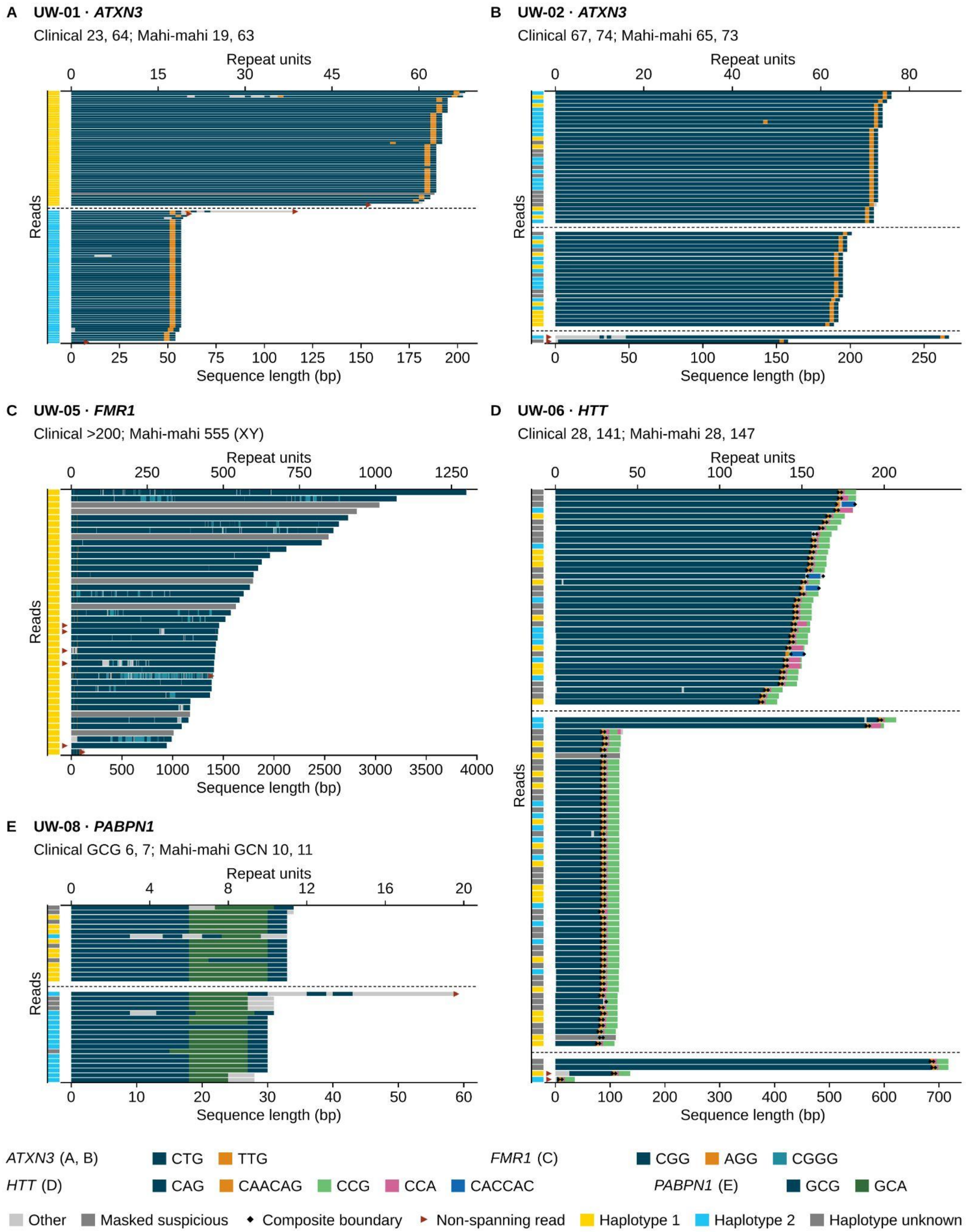

**Figure S10. Read-level repeat structure at additional loci in the UW cohort.**

Mahi-mahi read-level visualizations, displayed as in Figure 5. **(A, B)** *ATXN3* in UW-01 (orthogonal 23, 64; Mahi-mahi 19, 63) and UW-02 (orthogonal 67, 74; Mahi-mahi 65, 73), shown on the reverse strand, with CTG and TTG corresponding to CAG and CAA on the coding strand. **(C)** *FMR1* in UW-05, an XY sample with a full mutation reported by the lab as greater than 200 repeats and called by Mahi-mahi at 555. Reads span several hundred to more than 1,000 repeat units, illustrating the size heterogeneity of large expanded alleles; CGGG and AGG interruptions and masked reads are visible. **(D)** *HTT* in UW-06 (orthogonal 28, 141; Mahi-mahi 28, 147), showing the same composite structure as Figure 5C at a larger expansion. Reads are grouped by allele length, and the sidebar shows haplotype where assigned; reads not assigned to either allele are shown at the bottom. **(E)** *PABPN1* in UW-08, in which the GCN tract consists of GCG and GCA units. The referring laboratory reported GCG triplets (6, 7) whereas Mahi-mahi reports the full GCN tract (10, 11); the difference in reporting units is described in Methods.

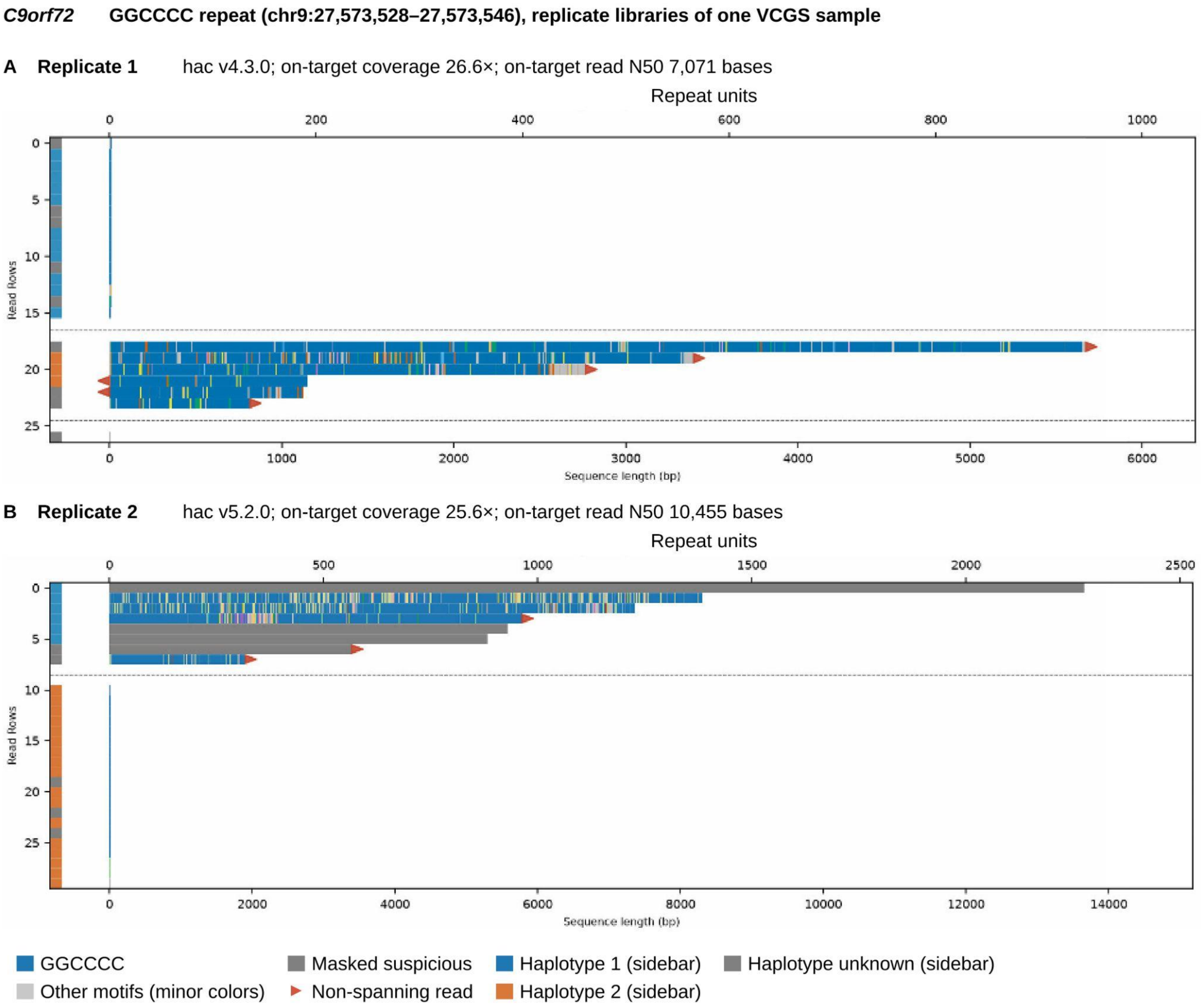

**Figure S11. Investigation of discrepancy between replicates of VCGS-27 at *C9orf72*.**

Mahi-mahi read-level visualizations, displayed as in Figure 5. **(A)** Replicate 1 (mean on-target coverage 26.6×; on-target read N50 7,071 bases; basecalled with the v4.3 HAC model). Every read from the expanded allele ends within the repeat (red triangles), so no read spans it, which likely accounts for the shorter repeat length called in this replicate (541 repeat units). **(B)** Replicate 2 (25.6×; N50 10,455 bases; v5.2 HAC). Three reads from the expanded allele are non-spanning and four are masked as suspicious (gray), but enough spanning reads remain to size the allele (1,208 repeat units) without using the non-spanning reads. Alleles are matched by length; haplotype labels differ between replicates.

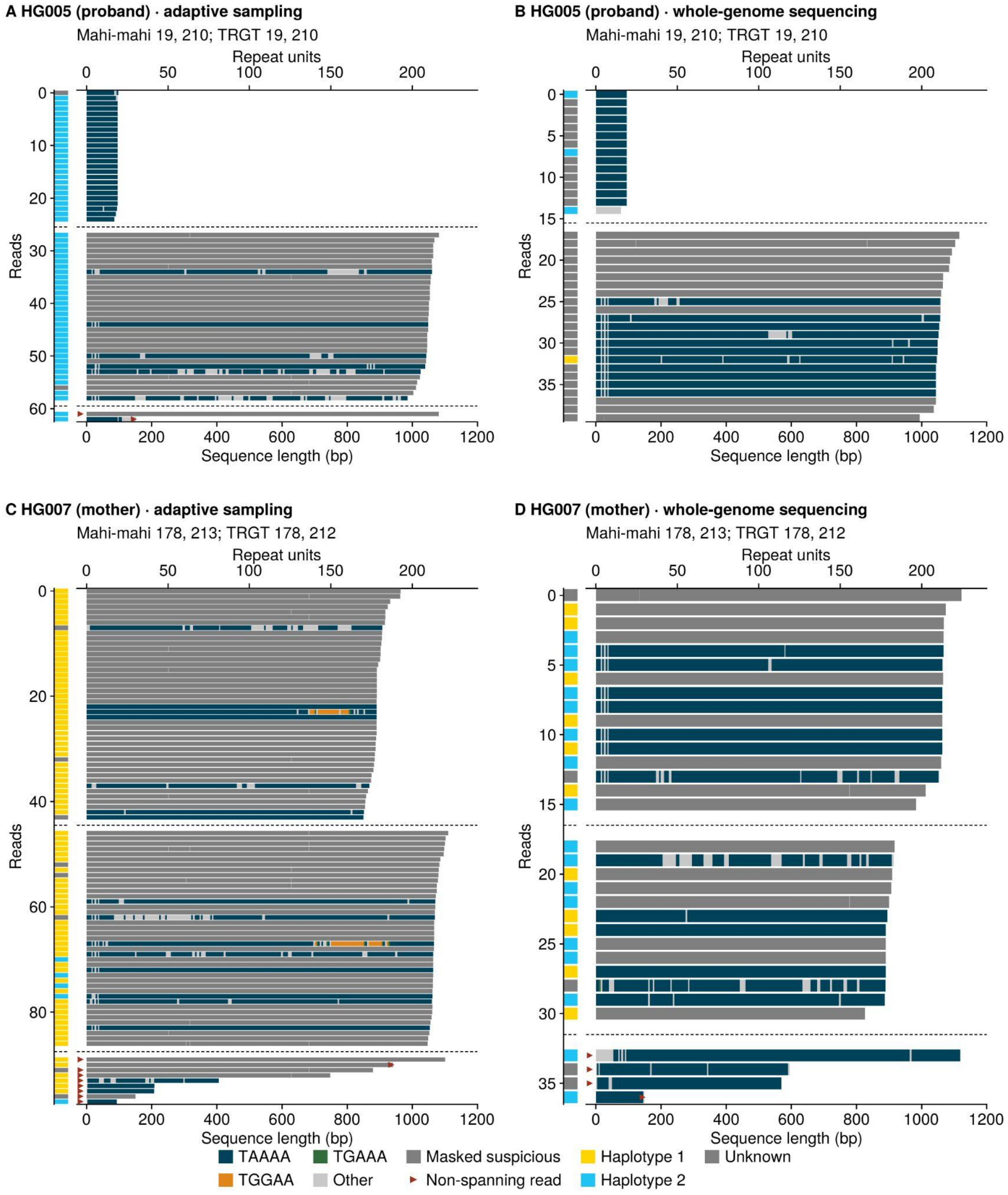

**Figure S12. *BEAN1* in the HG005 proband and his mother, HG007.**

Mahi-mahi read-level visualizations from adaptive sampling **(A, C)** and whole-genome sequencing **(B, D)** data. Each row is a read, reads are grouped by allele (dashed lines), the left sidebar indicates haplotype, colors indicate the motif assigned to each repeat unit, reads masked as suspicious are shown in dark gray, and red markers indicate reads that do not span the repeat. HG005 carries alleles of 19 and 210 repeat units, and HG007 carries two expanded alleles of 178 and 213 repeat units, of which the larger was transmitted to HG005. In both individuals most reads from the expanded alleles were masked as suspicious because of strand-biased miscalls, and the unmasked reads are composed of the canonical TAAAA motif; isolated TGGAA units appear in two HG007 reads. Mahi-mahi calls agree with TRGT to within one repeat unit in both individuals and with each other across sequencing strategies.
